# Innate and adaptive immune defects associated with lower SARS-CoV-2 BNT162b2 mRNA vaccine response in elderly people

**DOI:** 10.1101/2022.01.07.22268806

**Authors:** Joana Vitallé, Alberto Pérez-Gómez, Francisco José Ostos, Carmen Gasca-Capote, María Reyes Jiménez-León, Sara Bachiller, Inmaculada Rivas-Jeremías, Maria del Mar Silva-Sánchez, Anabel Ruiz-Mateos, Luis Fernando López-Cortes, Mohammed Rafii-El-Idrissi Benhnia, Ezequiel Ruiz-Mateos

**Affiliations:** Clinical Unit of Infectious Diseases, Microbiology and Preventive Medicine, Institute of Biomedicine of Seville (IBiS), Virgen del Rocío University Hospital, CSIC, University of Seville; Seville, 41013, Spain; Department of Medical Biochemistry, Molecular Biology, and Immunology, School of Medicine, University of Seville, Seville, 41009, Spain

**Keywords:** SARS-CoV-2, vaccine, BNT162b2 mRNA, elderly, immunosenescence, T cells, thymic function, dendritic cells, monocytes, inflammaging

## Abstract

The immune factors associated with impaired SARS-CoV-2 vaccine response in the elderly are mostly unknown. We studied old and young people vaccinated with SARS-CoV-2 BNT162b2 mRNA before and after the first and second dose. Aging was associated with a lower anti-RBD IgG levels and a decreased magnitude and polyfunctionality of SARS-CoV-2 specific T cell response. The dramatic decrease in thymic function in the elderly, which fueled alteration in T cell homeostasis, and lower CD161+ T cell levels were associated with decreased T cell response two months after vaccination. Additionally, a deficient dendritic cell (DC) homing, activation and Toll like receptor (TLR)-mediated function, along with a proinflammatory functional profile in monocytes, were observed in the elderly, which was also related to lower specific T cell response after vaccination. These findings might be relevant for the improvement of the current vaccination strategies and for the development of new vaccine prototypes.

## INTRODUCTION

Immune aging is sustained by multifaceted remodeling of the innate and adaptive immunity, which includes a diminished response to new antigens, a decreased memory T cell response and a persistent chronic inflammation (Aiello et al., 2019; Crooke et al., 2019; Franceschi et al., 2017; Pawelec, 2012). In consequence, immune aging leads to more severe consequences of viral infections as well as lower protection following vaccination (Connors et al., 2021). Severe acute syndrome coronavirus 2 (SARS-CoV-2) infection and its associated disease, COVID-19, are known to have a higher impact in old people. In fact, delayed viral clearance, prolonged disease and higher COVID-19 fatality rate have been related to age (Leung, 2020) and approximately 80% of hospitalizations involved people older than 65 years (Liu et al., 2020; Mueller et al., 2020).

Vaccination is the most effective tool for the prevention of the serious symptomatology caused by SARS-CoV-2 and other viral infections, especially for vulnerable populations as elderly people (Piot et al., 2019; Pulendran et al., 2010). BNT162b2 mRNA vaccine, commonly known as Biontech/Pfizer vaccine, has shown high safety and efficacy against severe outcomes of COVID-19 (Walsh et al., 2020). The two-dose vaccination of this SARS-CoV-2 vaccine induces a strong humoral response measured by the magnitude of binding antibodies to coronavirus Spike (S) protein and the neutralization capacity of the antibodies (Grigoryan and Pulendran, 2020; Walsh et al., 2020). In addition, notable SARS-CoV-2 S-specific CD4+ and CD8+ T cell responses have been observed after BNT162b2 vaccination (Agrati et al., 2021; Sahin et al., 2020; Zollner et al., 2021).

In spite of the promising results of the BNT162b2 vaccination, a lower effectiveness, in terms of COVID-19 symptoms, admissions to hospital and deaths after the vaccination, has been reported in elderly people (Haas et al., 2021; Lopez Bernal et al., 2021; Mazagatos et al., 2021). Moreover, other studies have described lower levels of neutralizing antibodies in vaccinated elderly comparing with younger subjects (Collier et al., 2021; Demaret et al., 2021; Müller et al., 2021), especially six months after the second dose (Levin et al., 2021; Tober-Lau et al., 2021). In addition, low levels of S-specific T cell response after vaccination have been shown in the elderly (Collier et al., 2021). However, the adaptive and innate immune factors associated with the lower vaccine response in elderly people are not yet characterized.

A better understanding of the age-related immune dysfunction to SARS-CoV-2 vaccine is crucial for future vaccination strategies to improve older adults’ protection against this virus. Thus, the aim of the study was to investigate the major immune alterations in old people, in terms of both SARS-CoV-2 specific adaptive and innate immunity, associated to a lower response to the SARS-CoV-2 BNT162b2 mRNA vaccine.

## RESULTS

### SARS-CoV-2 specific IgG levels are inversely associated with age

In this study, we included 54 healthy adults vaccinated with BNT162b2 mRNA vaccine against SARS-CoV-2, classified according to their age: 33 young people (median, 29 years [interquartile range, IQR 26-49]) and 21 elderly people (median 73 years [72-74]) (Figure S1). Three subjects were excluded from the study due to a positive result for SARS-CoV-2 RNA PCR or antibodies against receptor-binding domain (RBD) of the S protein of SARS-CoV-2 prior to vaccination. The innate and adaptive immunity parameters were analyzed before vaccination (PRE), three weeks after the first dose, just before the administration of the second dose (1D) and two months after the second dose (2D) (Figure S1).

Firstly, anti-RBD SARS-CoV-2 IgG levels were quantified by RBD-specific enzyme linked immunosorbent assay (ELISA) in old and young people at the three time points described above. In accordance to previous studies (Collier et al., 2021; Walsh et al., 2020), the BNT162b2 mRNA vaccine induced the production of SARS-CoV-2 specific IgG levels, being these levels much higher after the second dose comparing with the first dose (Figure 1A). Although no significant differences were observed between old and young subjects, young people tended to produce higher levels of specific antibodies after the first dose (Figure 1A). In fact, there was an inverse association between anti-RBD IgG levels and age after the administration of the first dose (Figure 1B, left), but not after the second one (Figure 1B, right). However, when we analyzed the young group alone, which presented a higher variability regarding to the age, we observed a negative correlation after both doses between anti-RBD IgG levels and age (Figure 1C).

**Figure 1.**
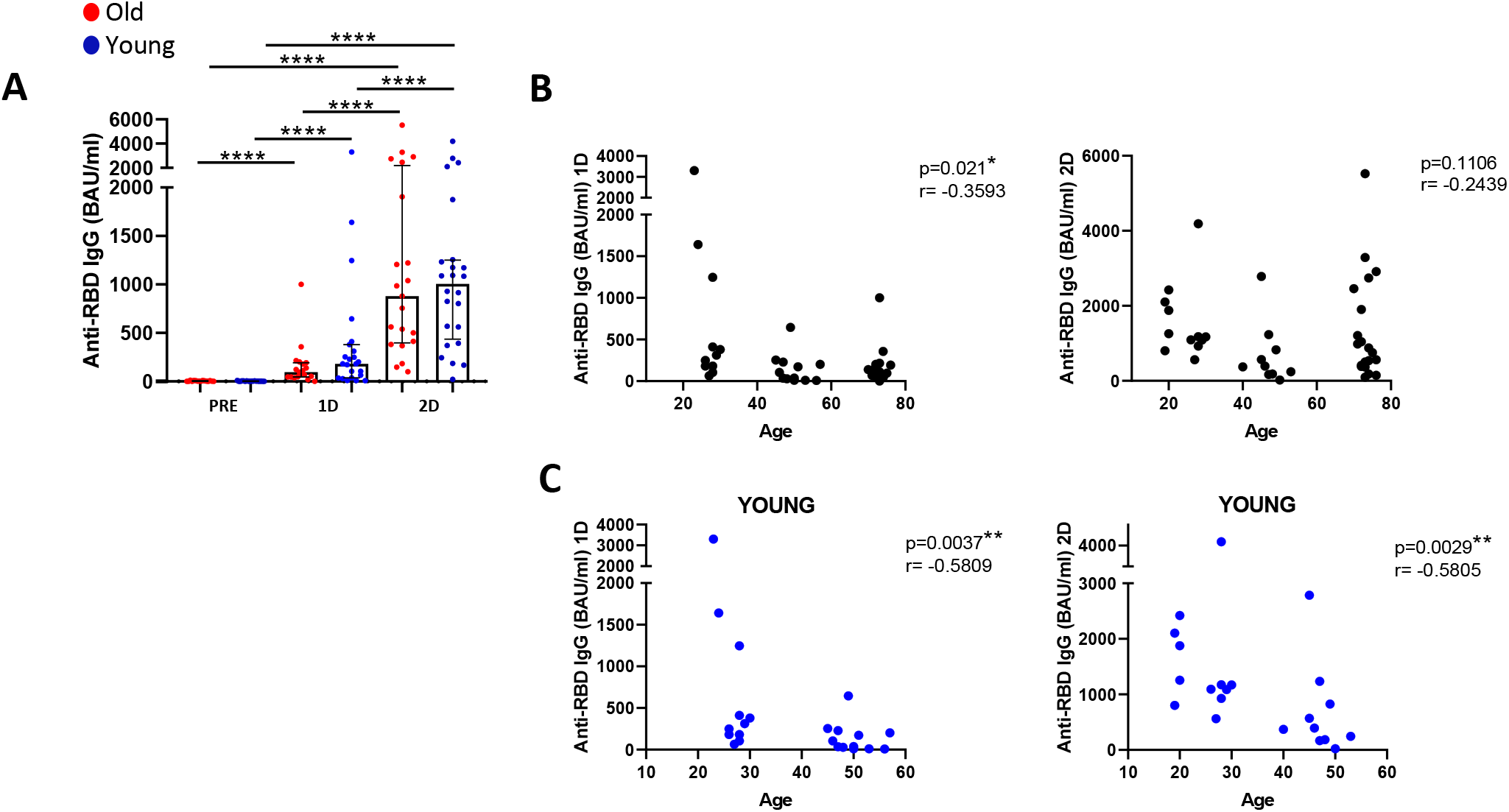
SARS-CoV-2 specific IgG levels are inversely associated with age. **(A)** Anti-RBD IgG levels (Binding Antibody Units (BAU)/mL) in old and young participants before SARS-CoV-2 vaccination (PRE), three weeks after the first dose (1D) and two months after the second dose (2D). **(B and C)** Correlation of anti-RBD IgG levels with age in all the study participants (B) and only in young group (C) after the first dose (left panels) and after the second dose (right panels).

### Old people show a lower and less polyfunctional SARS-CoV-2 Spike-specific CD4+ and CD8+ T cell response after vaccination

We investigated the magnitude and polyfunctionality of SARS-CoV-2 S-specific CD4+ and CD8+ T cell response through intracellular cytokine staining. We analyzed these parameters in total memory (Memory), central memory (CM), effector memory (EM) and terminal differentiated effector memory (TEMRA) CD4+ and CD8+ T cells (Figure S2A). Three weeks after the administration of the first dose, CD4+ T cells principally produced interferon gamma (IFN-γ) and tumor necrosis factor alpha (TNF-α) as an acute response to SARS-CoV-2 S protein, but also expressed low levels of the degranulation marker CD107a, perforin (PRF) and interleukin (IL)-2 (Figure S2B). Importantly, CD4+ T cells from elderly people showed a lower SARS-CoV-2 S-specific IFN-γ production and cytotoxic response, reflected in the percentage of CD107a+ and PRF+ cells, after the first dose of vaccination but mainly after the second dose (Figure 2A). In fact, the second dose of vaccination induced an increase in the cytotoxic function by CD4+ T cells in young people but not in old ones (Figure 2A, Figure S2C). Regarding CD8+, as it was observed in CD4+ T cells, elderly people showed a lower SARS-CoV-2 S-specific CD8+ T cell response, based mainly on the production of IFN-γ and the cytotoxic capacity (CD107a+ and PRF+), two months after the second dose (Figure 2B).

**Figure 2.**
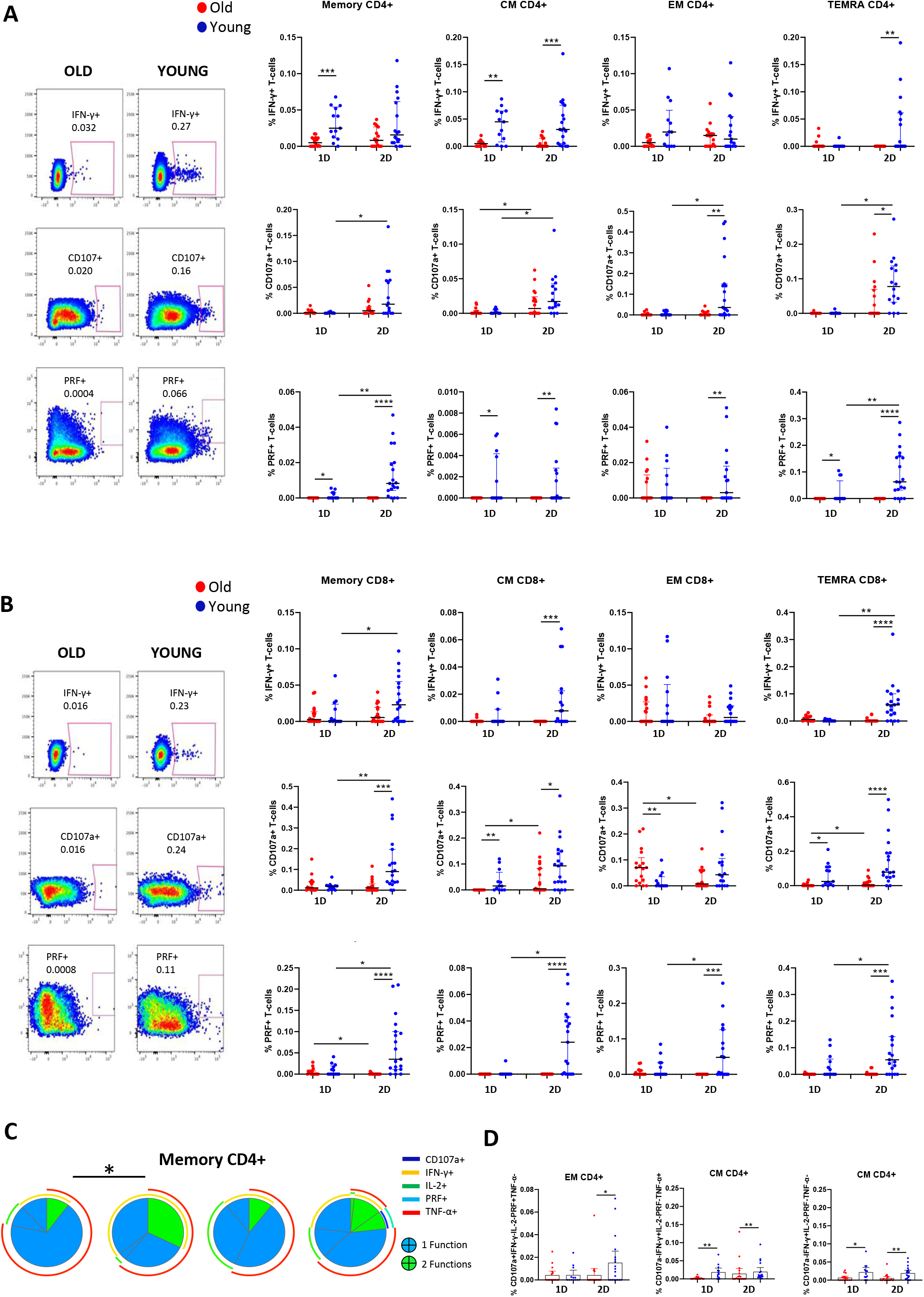
Old people show a lower and less polyfunctional SARS-CoV-2 spike-specific CD4+ and CD8+ T cell response after the vaccination. **(A and B)** Bar graphs presenting the percentage of IFN-γ+, CD107a+ and PRF+ cells within memory, CM, EM and TEMRA CD4+ (A) and CD8+ (B) T cells upon S-specific SARS-CoV-2 stimulation, comparing old and young subjects three weeks after the first dose (1D) and two months after the second dose (2D) of SARS-CoV-2 vaccine (right). Pseudocolour dot plot graphs show a representative data of memory CD4+ T cells from an old and young donor two months after vaccination (left). **(C)** Pie charts representing SARS-CoV-2 S-specific memory CD4+ T cell polyfunctionality. Each sector represents the proportion of S-specific CD4+ T cells producing two (green) or one (blue) functions. Arcs represent the type of function (CD107a, IFN-γ, IL-2, PRF and TNF-α) expressed in each sector. **(D)** Bar graphs showing the percentage of EM and CM CD4+ T cells expressing different combinations of studied functions (CD107a, IFN-γ, IL-2, PRF and TNF-α) comparing old and young subjects after the first (1D) and the second (2D) dose. See also Figure S2.

To determine the quality of the specific T cell response to SARS-CoV-2 vaccine, we analyzed the polyfunctionality of CD4+ and CD8+ T cells, which is defined by those cells that simultaneously produce multiple cytokines and degranulate (functions). In general, a low polyfunctional T cell response was observed after the vaccination with BNT162b2 mRNA vaccine in both old and young participants (Figure 2C). However, a more polyfunctional memory CD4+ T cell response to SARS-CoV-2 was observed in young people comparing with old ones after the first dose and a trend after the second dose (Figure 2C). The polyfunctional profile of the rest of CD4+ T cell subsets showed a similar pattern, with the exception of TEMRA CD4+ T cells that presented higher polyfunctionality after the second dose in young people (Figure S2D). To further characterize this SARS-CoV-2-specific T cell response to vaccination, we analyzed different combinations of the studied functions. CD4+ T cells expressing CD107a and PRF simultaneously were enriched in young people two months after the second dose of vaccination, and combinations including both IFN-γ and TNF-α and the ones including only IFN-γ were mainly observed in young people after the first and second dose of vaccination (Figure 2D). Furthermore, the percentages of T cells expressing three functions at the same time (e.g. IFN-γ+ CD107a+ PRF+ or IFN-γ+ IL-2+ TNF-α+) and other two-function combinations (e.g. IFN-γ+ IL-2+) were also higher in young than in old people (Figure S2E). Therefore, the SARS-CoV-2 S-specific T cell response is lower and less polyfunctional in elderly people after the vaccination with BNT162b2 mRNA vaccine.

### Lower thymic function and altered T cell homeostasis found in old people are associated to a lower response to the SARS-CoV-2 vaccine

Once we demonstrated that old people displayed a lower SARS-CoV-2 specific T cell response after vaccination, we investigated the immune defects that might be involved in the diminished response of this vulnerable population. In our group, we previously reported that thymic function failure and inflammation levels independently predict all-cause mortality in healthy elderly people (Ferrando-Martínez et al., 2013). Thus, we studied if these factors could be associated to a lower SARS-CoV-2 vaccine response in old subjects. Thymic output can be measured through the presence of T cell receptor rearrangement excision circles (TREC) in naïve T cells, indicators of recent thymic emigrants in humans (Douek et al., 1998). Thus, to determine the thymic function, we have developed an accurate and simplified technique using droplet digital PCR (ddPCR) which measure DβJβ-TREC and sj-TREC regions. Specifically, we analyzed the sj/β-TREC ratio that takes into account an intra-thymic proliferation step reflecting then thymus functionality, which is not altered by proliferation in the periphery (Dion et al., 2004, 2007). Our results showed a considerably lower level of thymic function in old people in comparison with young participants and accordingly, there was a strong association between thymic function and age (Figure 3A). In this line, we observed a correlation between thymic function with naïve CD4+ and CD8+ T cells, and with the naïve CD4+/CD8+ ratio (Figure 3B). In addition, decreased thymic function has been related to the phenomenon called *memory inflation*, which consists in the alteration of the naïve and memory T cell proportions in the periphery skewing toward memory T cells (Klenerman, 2018). This phenomenon was reflected in CD4+ and CD8+ T cell subsets distribution in old people, which displayed lower percentages of naïve T cells and higher percentages of memory T cells than young participants (Figure 3C). Three weeks after the vaccination with the first dose, young subjects showed a trend to decrease the percentages of naïve T cells and to increase memory T cells and a restoration of T cell subset distribution occurred two months after the second dose (Figure 3C). Thus, the differences in naïve and memory T cells between old and young people observed prior to vaccination were lost after the first dose but were restored two months after the second dose (Figure 3C).

**Figure 3.**
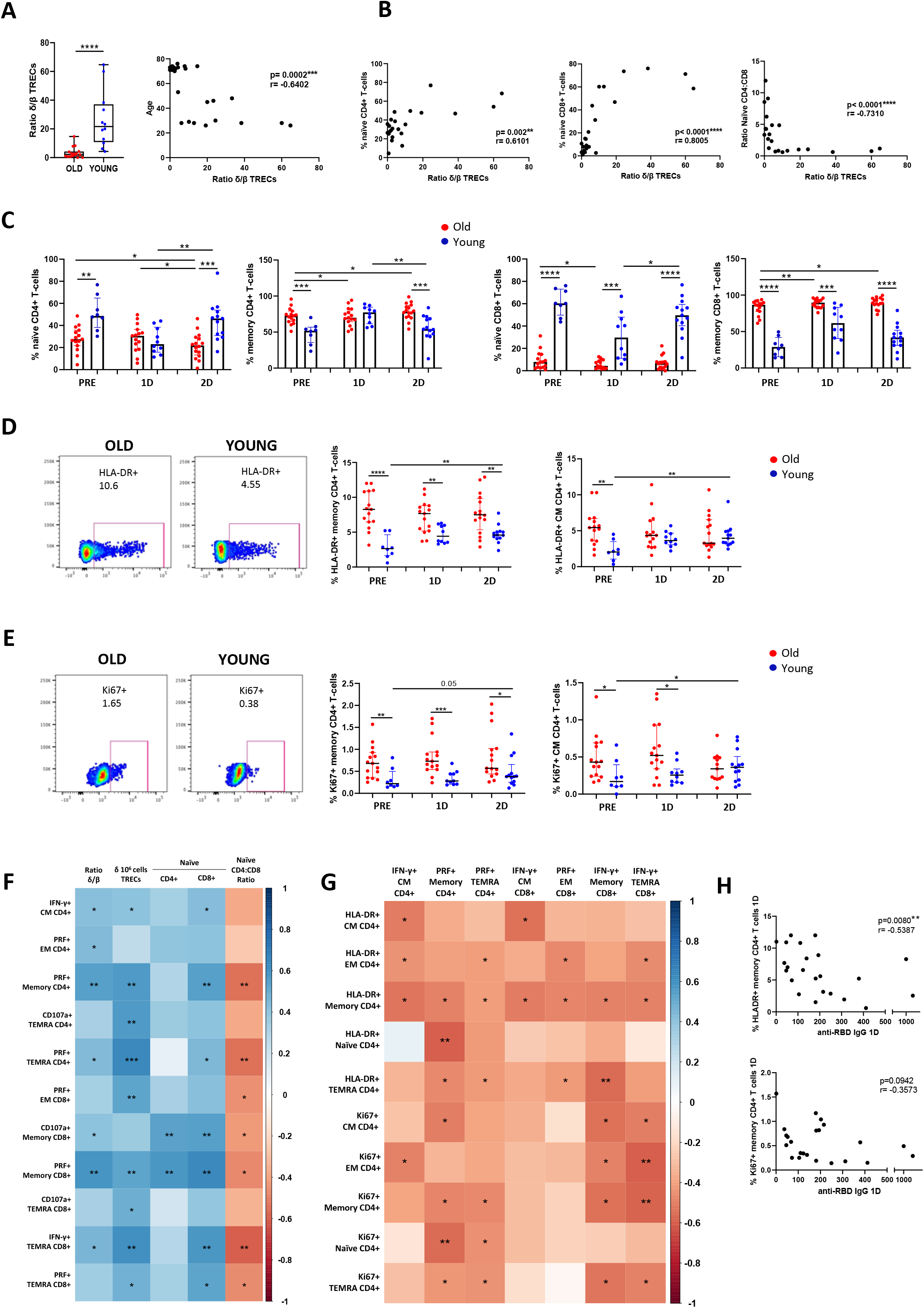
Lower thymic function and altered T cell homeostasis found in old people are associated to a lower response to the SARS-CoV-2 vaccine. **(A and B)** Bar graphs showing sj/β-TREC ratio as a measure of thymic function in old and young participants prior to vaccination (A, left) and the correlation of the sj/β-TREC ratio with age (A, right), naïve CD4+ T cells (B, left), naïve CD8+ T cells (B, middle) and naïve CD4+/CD8+ T cell ratio (B, right). (C) Bar graphs representing the percentage of naïve and memory CD4+ and CD8+ T cells in old and young participants before SARS-CoV-2 vaccination (PRE), three weeks after the first dose (1D) and two months after the second dose (2D). **(D and E)** Bar graphs representing the percentage of memory and CM CD4+ T cells expressing HLA-DR (D) and Ki67 (E) in old and young participants at the three time points (right). Pseudocolour dot plot graphs show representative data of memory CD4+ T cells from an old and young donor expressing HLA-DR (D) and Ki67 (E) before vaccination (left). **(F and G)** Correlation matrixes representing associations of SARS-CoV-2 S-specific CD4+ and CD8+ T cells expressing IFN-γ or cytotoxicity markers two months after the second dose of vaccination with sj/β-TREC ratio, DβJβ-TREC/106 cells and naïve T cells (F) and with the percentage of HLA-DR+ and Ki67+ CD4+ T cells (G) before vaccination in all participants. **(H)** Correlation plots of anti-RBD IgG levels after the first dose of vaccination with the percentage of HLA-DR+ and Ki67+ CD4+ T cells before vaccination. See also Figure S3.

According to lower thymic function and high memory inflation, elevated levels of T cell homeostatic proliferation and activation were found in the elderly people. Specifically, we observed that prior to vaccination, old people displayed a higher percentage of activated (HLA-DR+) and proliferating (Ki67+) memory and CM CD4+ T cells comparing with young people (Figure 3D and 3E). After vaccination with two doses, an increase of HLA-DR+ and Ki67+ CD4+ T cells was observed in young subjects, while older people showed a lack of further activation and proliferation through vaccination (Figure 3D and 3E). Similar results were found in the rest of CD4+ T cell subsets (Figure S3A and S3B) and in some of the CD8+ T cell subsets (data not shown). In this line, thymic function was negatively correlated with T cell activation in most of the subsets and proliferation in naïve CD8+ T cells (Figure S3C). Moreover, the vaccination altered the expression of immune checkpoints as LAG-3, PD-1 and TIGIT on CD4+ and CD8+ T cells being higher in young compared with old people after the first or second dose in most of the T cell subsets (Figure S3D, S3E and S3F). Importantly, thymic dysfunction and the related defects in the homeostasis of T cell compartment found in elderly people prior to vaccination, expressed as higher T cell activation and proliferation, along with memory inflation, were correlated with a lower SARS-CoV-2-specific CD4+ and CD8+ T cell response after the two-dose vaccination (Figure 3F-G). Furthermore, the percentage of activated (HLA-DR+) memory CD4+ T cells was also inversely associated to anti-RBD IgG levels after the first dose and the same trend was observed regarding proliferating (Ki67+) T cells (Figure 3H). In addition, virus specific T cell response after vaccination was also inversely correlated with the expression of PD-1 before vaccination (Figure S3G).

Other T cells that notably differed between old and young subjects were CD161+ T cells. These cells mainly produce IL-17 and are known to have an important contribution in pathogen clearance (Dominguez-Molina et al., 2018). CD161+ T cells presented higher levels in young people comparing with old ones, independently on the vaccination (Figure 4A). It is remarkable, that the percentage of CD161 expressing CD4+ and CD8+ T cells was positively associated to SARS-CoV-2 S-specific T cell response two months after the vaccination (Figure 4B).

**Figure 4.**
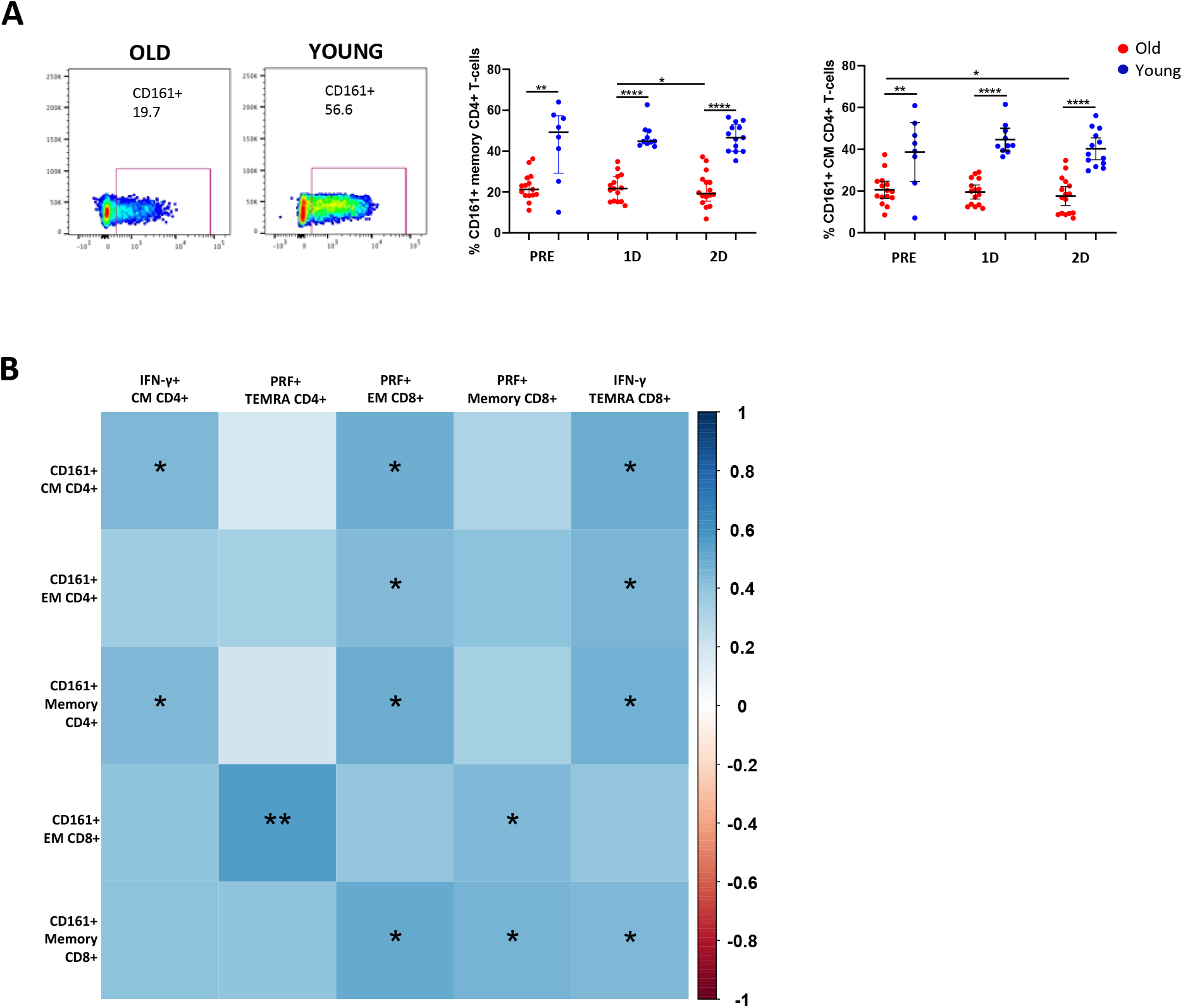
CD161 expressing T cell levels are associated to a higher SARS-CoV-2 vaccine response. **(A)** Bar graphs representing the percentage of memory and CM CD4+ T cells expressing CD161 in old and young participants before SARS-CoV-2 vaccination (PRE), three weeks after the first dose (1D) and two months after the second dose (2D) (right). Pseudocolour dot plot graphs show representative data of memory CD4+ T cells from a young and old donor expressing CD161 before vaccination (left). **(B)** Correlation matrix representing associations of the percentage of CD161+ T cells before vaccination with SARS-CoV-2 S-specific CD4+ and CD8+ T cells expressing IFN-γ or cytotoxicity markers two months after the second dose of vaccination in all participants.

### An impaired dendritic cells homing and functional capacity are associated to a lower response to SARS-CoV-2 vaccine in old people

In addition to the alteration of the adaptive immunity associated with lower response to the vaccine in the elderly, there is a remodeling of the innate immune system with aging (Shaw et al., 2010). Thus, we next studied plasmacytoid dendritic cells (pDCs) and myeloid dendritic cells (mDCs) (Figure S4A), innate immune cells with a key role in the modulation of T cell response (Guermonprez et al., 2002). We first observed a decrease in pDC percentages two months after the second dose (Figure 5A). Although we did not find differences between young and elderly people in pDC levels (Figure 5A), we observed a considerable difference in pDC functional capacity (Figure 5B). When cells were stimulated in a toll like receptor (TLR)-9-dependent manner by CpG-A, a lower IFN-α production was observed in elderly people comparing with young subjects, both after the first and second dose of the SARS-CoV-2 vaccine (Figure 5B, left panel). Interestingly, we also observed how this functional capacity of the pDCs was associated with anti-RBD IgG levels after the first dose (Figure 5B, right panel).

**Figure 5.**
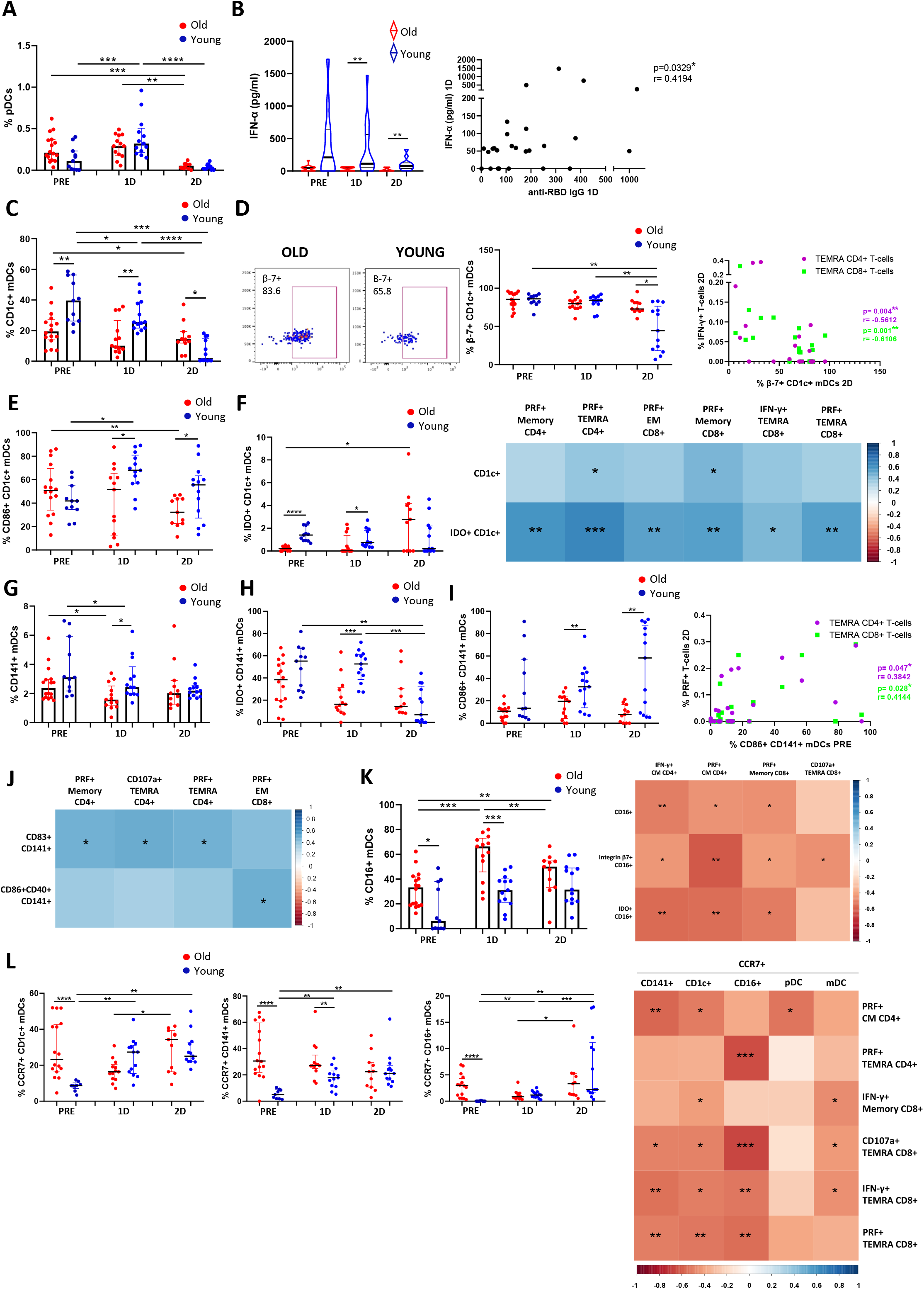
An impaired DC homing and functional capacity are associated to a lower response to SARS-CoV-2 vaccine in old people. **(A)** Bar graphs representing the percentage of pDCs in old and young participants before SARS-CoV-2 vaccination (PRE), three weeks after the first dose (1D) and two months after the second dose (2D) of vaccination. **(B)** Violin plots showing IFN-α production through CpG-A stimulation for 18 hours in old and young participants before SARS-CoV-2 vaccination (PRE), three weeks after the first dose (1D) and two months after the second dose (2D) (left). Correlation analysis of IFN-α production with anti-RBD IgG levels three weeks after the first dose of vaccination (right). **(C)** Bar graphs representing the percentage of CD1c+ mDCs in old and young participants at the three follow up time points. **(D)** Dot plots showing the percentage of integrin-β7+ CD1c+ mDCs in old and young participants: pseudocolour plots show a representative data of a young and old participants two months after the second dose (left) and bar graphs show data from all subjects at the three time points (middle). Correlation plot between the percentage of integrin-β7+ CD1c+ mDCs and the percentage of S-specific IFN-γ+ TEMRA CD4+ and CD8+ T cells two months after the second dose (right). **(E and F)** Dot plots showing the percentage of CD1c+ mDCs expressing CD86 (E), and IDO (F, left) in old and young participants at the three time points. Correlation matrix showing associations between the percentages of CD1c+ mDCs and IDO+ CD1c+ mDCs before vaccination with SARS-CoV-2 S-specific T cells expressing cytokines or cytotoxicity markers two months after the second dose (F, right). **(G-I)** Bar and dot plot graphs showing the percentage of CD141+ mDCs (G) and CD141+ mDCs expressing IDO (H), and CD86 (I, left) in old and young participants at the three time points. Correlation plot between the percentage of CD86+ CD141+ mDCs before vaccination and the percentage of S-specific PRF+ TEMRA CD4+ and CD8+ T cells two months after the second dose (I, right). **(J)** Correlation matrix showing associations between the percentage of CD141+ mDCs expressing activation markers after TLR-3 stimulation for 24 hours with SARS-CoV-2 S-specific CD4+ and CD8+ T cells expressing cytotoxicity markers. **(K)** Bar graphs representing the percentage of CD16+ mDCs in old and young participants at the three time points (left). Correlation matrix showing associations between the percentage of CD16+ mDCs and CD16+ mDCs expressing integrin-β7 and IDO with SARS-CoV-2 S-specific CD4+ and CD8+ T cells expressing cytotoxicity markers (right). **(L)** Dot plots showing the percentage of CCR7+ mDCs in old and young participants in the three follow up time points (left and middle panels) and correlation matrix representing associations of the percentage of mDCs expressing CCR7 with SARS-CoV-2 S-specific CD4+ and CD8+ T cells expressing cytokines or cytotoxicity markers two months after the second dose (right). See also Figure S4.

Next, we focused on mDC subsets, including CD1c+, CD16+ and CD141+ mDCs (Figure S4A). Our results showed a higher percentage of CD1c+ mDCs in young than in old people prior to vaccination and after the first dose (Figure 5C), a mDC subpopulation that modulates CD4+ T cell response (Guermonprez et al., 2002). In contrast, a notable decrease in CD1c+ mDC percentage was observed only in young people two months after de second dose of the SARS-CoV-2 vaccine (Figure 5C). The same result was found regarding the percentage of integrin-β7 expressing CD1c+ mDCs (Figure 5D, left and middle panel), being integrin-β7 a maker of cell homing to gut. It is remarkable, that the decrease in integrin-β7+ CD1c+ mDCs was correlated with a higher IFN-γ production by TEMRA CD4+ and CD8+ T cells in response to SARS-CoV-2 two months after vaccination (Figure 5D, right). Furthermore, higher expression of the activation markers CD86 and Indoleamine 2,3-dioxygenase (IDO) was found on CD1c+ mDCs from young people in most of the studied time points (Figure 5E and 5F left panel). In fact, the percentage of CD1c+ and IDO+ CD1c+ mDCs prior to vaccination were correlated with SARS-CoV-2-specific T cell response two months after the second dose (Figure 5F right panel).

It is also known that CD141+ mDCs are involved in the regulation of CD8+ T cell response (Guermonprez et al., 2002). In this study, higher CD141+ mDC levels were found in young people after the first dose, comparing with old ones (Figure 5G). As occurred with CD1c+ mDCs, young people also displayed a higher expression of the activation markers IDO (Figure 5H) and CD86 (Figure 5I, left panel) after vaccination in this subset and this was correlated with SARS-CoV-2 specific cytotoxic response (PRF+) by TEMRA T cells (Figure 5I, right panel). In order to study the functional capacity of mDCs, cells were stimulated with Poly I:C, an agonist of TLR-3. In all subjects, the percentages of activated CD141+ mDCs (CD86+CD40+ and CD83+) were increased following TLR-3-stimulation, before vaccination and after the first dose (Figure S4B). Nevertheless, two months after the second dose, CD141+ mDCs were not successfully stimulated via TLR-3, since they were already activated by the vaccination (Figure S4B). This effect was also observed in CD1c+ mDCs but not in CD16+ mDCs (Figure S4C). Even though no differences were found between old and young people in CD141+ mDC response, a higher functional capacity of CD141+ mDCs prior to vaccination was positively associated with SARS-CoV-2 specific T cell response after vaccination (Figure 5J).

Lastly, due to their role in the modulation of inflammatory responses, CD16+ mDCs were also studied (Piccioli et al., 2007). In general, elderly people displayed higher percentages of CD16+ mDCs than young subjects (Figure 5K, left panel). Focusing on the effect of SARS-CoV-2 vaccination, an increase in CD16+ mDC percentages were observed mainly after the first dose but also after the second dose only in old subjects (Figure 5K, left panel). The percentage of CD16+ mDCs and the percentages of CD16+ mDCs expressing integrin β7 and IDO were inversely associated with S-specific T cell response (Figure 5K, right panel). Additionally, other DC markers were altered after the vaccination on different subsets. CCR7, a chemokine receptor that orchestrates DC migration to draining lymph nodes (Rodríguez-Fernández and Criado-García, 2020), was highly expressed in old people before vaccination and its expression was increased after the first and second dose mostly in young people, with no response in elderly people (Figure 5L, left and middle panels). A higher CCR7 expression on DCs prior to vaccination was associated to a lower SARS-CoV-2 specific T cell response (Figure 5L, right panel). The receptors PDL-1 and CD4 were also overexpressed on several DC subsets after the SARS-CoV-2 vaccination and specially after the first dose, this upregulation was observed in young but not in old people (Figure S4E and S4F).

Hence, our results indicate that the higher levels of CD16+ mDCs, which have a pro-inflammatory function, along with the impaired DC homing and functional capacity found in elderly people were associated to a lower T cell response to SARS-CoV-2 vaccine.

### Differential phenotypical and functional pattern in monocytes from elderly people is related to a lower response to the SARS-CoV-2 vaccine

As one of the main players in inflammatory responses and the inflammaging phenomenon (De Maeyer and Chambers, 2021; Shaw et al., 2010), monocytes were analyzed in this study. Specifically, classical (CD14++ CD16), intermediate (CD14++ CD16+) and non-classical monocytes were studied (CD14+ CD16++) (Figure S5A). Our results showed that in all subjects, SARS-CoV-2 vaccination induced an increase in the expression of activation markers in classical and intermediate monocytes, including CD40, TLR-4 (Figure 6A, left panel and Figure S5B), TLR-2 and CD49d (Figure S5C and S5D). Nevertheless, the monocyte activation levels after the first dose were higher in young people than elderly people (Figure 6A, left panel and Figure S5B). It is remarkable, that the expression of CD40 and TLR-4 before vaccination were positively correlated to SARS-CoV-2-specific T cell response two months after the second dose (Figure 6A, right panel). The expression of CCR5, a monocyte chemokine receptor, is modulated after activation (Oppermann, 2004). Our results showed that CCR5 expression before vaccination was higher in monocytes from young than in old people (Figure 6B, left panel). However, it was downregulated after the first dose of SARS-CoV-2 vaccination, being this decrease less pronounced in old people (Figure 6B, middle panel). Importantly, basal CCR5 expression was also directly correlated to IFN-γ production by CM CD4+ and TEMRA CD8+ T cells after SARS-CoV-2 vaccination (Figure 6B, right panels). Regarding other monocyte chemokine receptors, two-dose vaccination induced a decrease in the percentage of intermediate monocytes expressing CCR2 and CD11b and non-classical monocytes expressing CX3CR1, being these percentages lower in young people than in old ones (Figure 6C-E, left panels). The lower percentages of monocytes expressing these chemokine receptors two months after the second dose of vaccination was inversely correlated with SARS-CoV-2 T cell response and regarding CD11b, also with anti-RBD IgG levels (Figure 6C-E, right panels). Furthermore, the expression of CX3CR1 in non-classical monocytes prior to vaccination was also inversely associated to the virus-specific T cell response (Figure 6F). Moreover, monocyte tissue factor (CD142) expression is known to be induced by several inflammatory stimuli (Kothari et al., 2012; Levi et al., 2004). Here, we found an increase in the expression of tissue factor on classical monocytes after SARS-CoV-2 vaccination in both groups (Figure S5E, left panel). In addition, tissue factor expression levels were higher on intermediate monocytes from young subjects after the first dose (Figure S5E).

**Figure 6.**
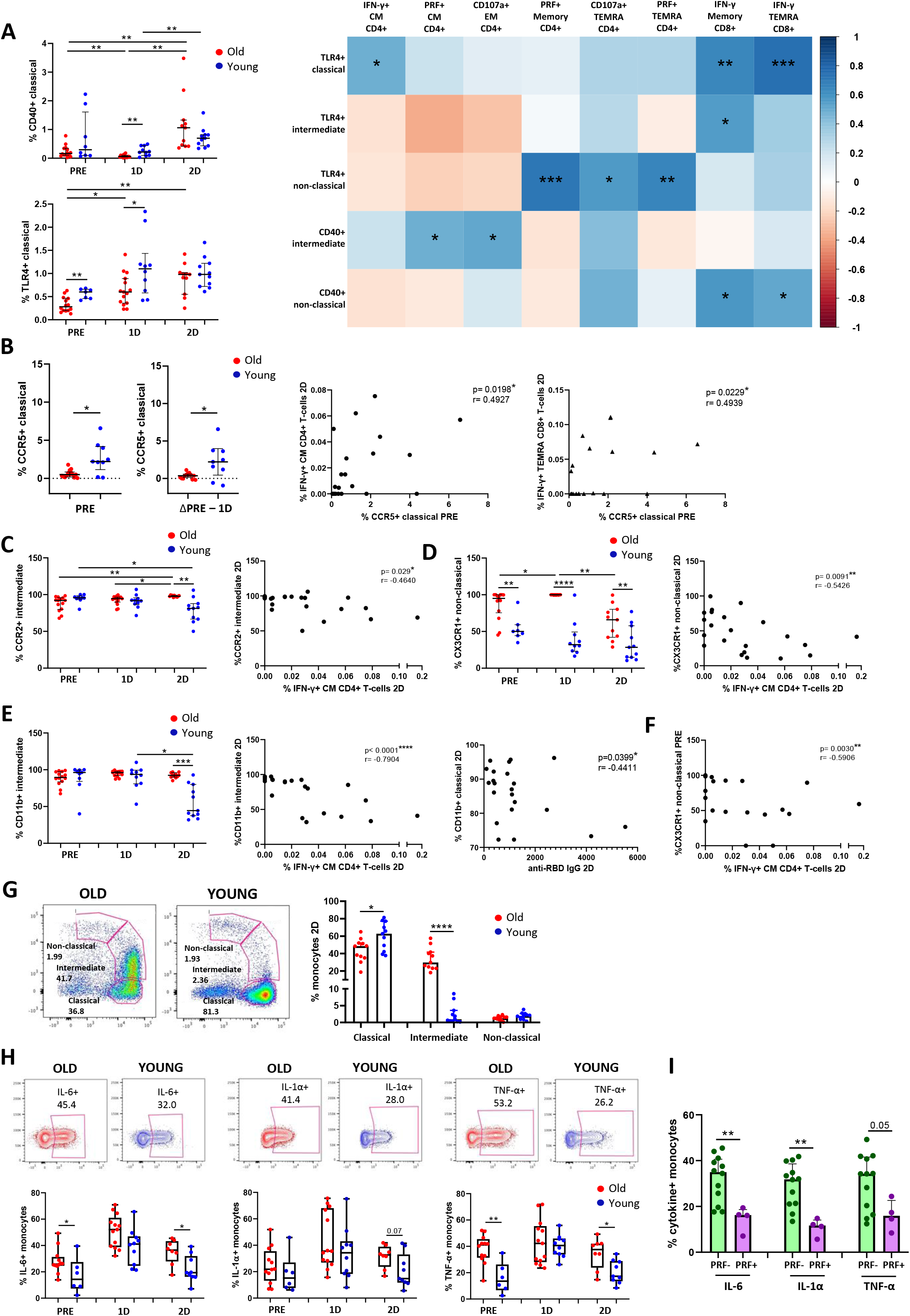
Differential phenotypical and functional pattern in monocytes from elderly people is related to a lower response to the SARS-CoV-2 vaccine. **(A)** Dot plots representing the percentage of classical monocytes expressing CD40 and TLR-4 in old and young participants before SARS-CoV-2 vaccination (PRE), three weeks after the first dose (1D) and two months after the second dose (2D) of vaccination (left). Correlation matrix representing the percentage of monocytes expressing CD40 and TLR-4 prior to vaccination with SARS-CoV-2 S-specific CD4+ and CD8+ T cells expressing cytokines or cytotoxicity markers two months after the second dose (right). (B) Dot plots representing the percentage of classical monocytes expressing CCR5 before vaccination (left), the fold of decrease in the percentage of CCR5+ cells after the first dose (middle) and correlation analysis of the percentage of CCR5+ classical monocytes prior to vaccination with SARS-CoV-2 S-specific CD4+ and CD8+ T cells expressing IFN-γ two months after the second dose (right). **(C-E)** Dot plots showing the percentages of monocytes expressing CCR2 (C), CX3CR1 (D) and CD11b (E) in old and young subjects at the three follow up time points (left) and correlations of the percentage of monocytes expressing these markers with SARS-CoV-2 S-specific IFN-γ+ CD4+ T cells two months after the second dose. Correlation plot between the percentage of CD11b+ classical monocytes with anti-RBD IgG levels two months after the second dose (E, right). **(F)** Correlation analysis of the percentage of CX3CR1+ non-classical monocytes before vaccination with SARS-CoV-2 S-specific IFN-γ+ CM CD4+ T cells after the second dose. **(G)** Bar graphs representing the percentage of classical, intermediate and non-classical monocytes in old and young participants at the three time points (right). Pseudocolour plots showing representative data of an old and young participant two months after the second dose (left). **(H)** Box and whiskers graphs representing the percentage of IL-6+, IL-1α+ and TNF-α+ monocytes upon TLR-4 stimulation in old and young participants at the three time points (bottom). Contour plots showing representative data of the percentage of cytokine+ monocytes from an old and young subject two months after the second dose (top). **(I)** Bar graphs showing the percentage of cytokine (IL-6, IL1-α and TNF-α) producing monocytes on individuals with a cytotoxic SARS-CoV-2 S-specific T cell response and the ones with a negative response. The percentage of specific PRF+ T cells higher than 0.01 was considered as a positive cytotoxic T cell response. See also Figure S5.

It has been previously reported that intermediate monocytes are expanded in the blood of patients with systemic infections (Kapellos et al., 2019). Here, we found a considerable increase in the percentage of intermediate monocytes, along with a decrease in classical monocytes, after the two-dose SARS-CoV-2 vaccination only in old people (Figure 6G). Intermediate monocytes are known to secrete TNF-α, IL-1β, IL-6, and CCL3 upon TLR stimulation (Kapellos et al., 2019). In this study, we stimulated cells in a TLR-4 dependent manner by adding lipopolysaccharide (LPS). Importantly, vaccinated elderly people showed a higher production of IL-6, IL-1α and TNF-α by monocytes upon LPS stimulation comparing with young subjects, mainly after the second dose (Figure 6H). Lastly, we discovered that participants which did not show a cytotoxic SARS-CoV-2 specific T cell response (PRF-) produced higher levels of inflammatory cytokines by monocytes after vaccination, than the ones presenting cytotoxic T cells (PRF+) after SARS-CoV-2 vaccination (Figure 6I). In summary, although monocytes from old people showed lower levels of activation and homing after the vaccination, produced higher levels of proinflammatory cytokines upon LPS stimulation which was inversely associated to SARS-CoV-2 specific T cell response.

## DISCUSSION

Aging is associated with impaired COVID-19 vaccine response (Collier et al., 2021). We confirmed and extended that the age-related immunological defects were characterized by lower anti-RBD IgG levels and mostly by a lower magnitude and polyfunctionality of SARS-CoV-2 specific T cell response. The adaptive and innate immune factors behind these defects in elderly people included an alteration in T cell homeostasis parameters fueled by lower thymic function and higher T cell activation and proliferation, dendritic cell dysfunction and a higher proinflammatory profile in circulating monocytes.

In spite of the efficacy of BNT162b2 mRNA vaccine to prevent severe COVID-19 outcomes, vulnerable populations as elderly people remain at risk. Several studies have reported lower humoral response in old subjects following vaccination (Collier et al., 2021; Demaret et al., 2021; Müller et al., 2021) (Levin et al., 2021; Tober-Lau et al., 2021). In accordance to these findings, we observed an inverse association between anti-RBD IgG levels with age two months after vaccination. Moreover, we also found a diminished specific T cell response to SARS-CoV-2 vaccine in old people, confirming previous results (Collier et al., 2021; Demaret et al., 2021). In addition to the magnitude, high quality of SARS-CoV-2 specific T cell function is required to achieve an effective vaccine response, which has not been completely determined yet. We found that specific T cells in addition to produce different cytokines, also exhibited a cytotoxic response to the vaccination, which is diminished in elderly. Although young people showed a higher polyfunctional response than old ones, BNT162b2 mRNA vaccine did not induce a high polyfunctional T cell immunity in general. This could be one of the key factors that might explain the absence of vaccine efficacy to avoid new infections along time, independently of the virus variant of concern (VOC).

Age-related changes in the immune system, known as immunosenescence, cause a subclinical immune deficiency that involves a reduced antiviral function and vaccine response (Connors et al., 2021). One of the most known changes of the immune aging is the involution of the thymus (Palmer, 2013) (Klenerman, 2018). In our study, old people exhibited thymic dysfunction and the subsequent memory inflation, which was associated to a higher homeostatic T cell proliferation and activation, reproducing previous findings (Ferrando-Martínez et al., 2011). In fact, the age-dependent shift in the T cell population from naïve to memory phenotype induces homeostatic T cell proliferation to compensate the diminished T cell thymic output (Kilpatrick et al., 2008; Sato et al., 2017). Remarkably, it has been described that thymic function failure predicts all-cause mortality in healthy elderly people (Ferrando-Martínez et al., 2013) and has a relevant role in viral infections such as HIV-1 infection (Delgado et al., 2002; Douek et al., 1998; Ferrando-Martinez et al., 2017). Moreover, it has been suggested that thymic aging might have an important implication in COVID-19 disease severity (Kellogg and Equils, 2021; Wang et al., 2021). Importantly, we discovered that the thymic dysfunction, along with the memory inflation and a higher homeostatic T cell proliferation and activation found in old people, were correlated to a lower response to SARS-CoV-2 vaccination. Accordingly, a lower activity of the thymus has been previously associated to diminished responsiveness to vaccination against other viruses as yellow fever virus (Schulz et al., 2015).

One of the most remarkable findings of our study is the tight association of CD161 expression levels with age. CD161 is a C-type lectin receptor which is expressed in both T and NK cells (Lanier et al., 1994). CD161+ T cells has been associated with IL-17 production and hence with pathogen clearance (Merino et al., 2021). In fact, we have previously observed that CD161+ CD8+ T cells were related with HIV and hepatitis C virus (HCV)-specific T cells polyfunctionality, which is essential for HIV spontaneous control and HCV spontaneous clearance (Dominguez-Molina et al., 2018). Results presented herein suggest CD161 marker as a hallmark of T cell immunosenescence in accordance with recent findings, which show a correlation of CD161 expression on CD8+ T cells with age independently of CMV infection (Formentini et al., 2021). Importantly, a reduction in the frequency of CD161+ CD8+ T cells was found in peripheral blood of severe COVID-19 patients (Kuri-Cervantes et al., 2020). In this study, we found CD161 expression on T cells was tightly associated with SARS-CoV-2 vaccine response, highlighting this molecule as a potential target for immunotherapeutic strategies for age-related disease therapies and vaccine response in the elderly.

The regulation of the immune response highly depends on the function of DCs (Guermonprez et al., 2002); however, their role in immune aging needs to be better understood. In this work, we found an altered DC subset distribution and an impaired DC homing and function with aging. The levels of CD1c+ mDCs, cells responsible for the modulation of CD4+ T cell response, were lower in old people prior to vaccination and this was related to a lower SARS-CoV-2 T cell response after vaccination. Accordingly, Schulz et al. previously observed an association between DC numbers and the response to yellow fever vaccine (Schulz et al., 2015). Furthermore, the lower numbers of CD1c+ mDCs and integrin-β7 expressing CD1c+ mDCs observed after the second dose indicates that vaccination may induce a CD1c+ mDC migration to gut and probably to other tissues. In this line, a decrease in peripheral blood CD1c+ mDC numbers has been found in patients with severe COVID-19, due to a migration of these cells from blood to the lungs (Pérez-Gómez et al., 2021; Sánchez-Cerrillo et al., 2020). Interestingly, the vaccine-induced CD1c+ homing was barely observed in old people and this was also associated to a decreased SARS-CoV-2 specific T cell response, suggesting that aging might affect the capacity of CD1c+ mDCs to migrate and consequently also their ability to modulate T cell function.

In addition to homing, DC function is also altered with aging. In response to TLR-3, TLR-7/8, and TLR-9 ligand stimulation, CD141+ mDCs and pDCs from old subjects secreted lower levels of IL-6, IL-12, and TNF-α (Panda et al., 2010). Other studies have reported, that in response to influenza A virus infection and West Nile infection, pDCs from older adults produced less type I IFN (Prakash et al., 2013). Accordingly, our results showed a lower TLR-9-dependent IFN-α production by pDCs in elderly people, both before and after the vaccination against SARS-CoV-2. According to this, patients with COVID-19 presented a lower TLR-9-mediated IFN-α production than healthy donors (Pérez-Gómez et al., 2021). This might be of great importance, since type I IFNs control innate and adaptive immune system and induce cells’ antiviral state via the upregulation of IFN-stimulated genes that inhibit the replication and spreading of viruses (Bencze et al., 2021). Remarkably, we described that CD141+ mDCs may also have an important implication in SARS-CoV-2 vaccination immunity, which is one of the cell types that is known to be depleted in COVID-19 patients and are important for disease progression (Huang et al., 2021; Sánchez-Cerrillo et al., 2020). Here, we discovered that TLR-3-mediated CD141+ mDC activation capacity was directly associated to SARS-CoV-2 T cell response following vaccination. This is in accordance with the important function of CD141+ mDCs in antigen presentation to T cells (Collin and Bigley, 2018; Macri et al., 2018).

Other important innate immune cells that might have a relevant role in vaccine response are monocytes. In this study, we reported that SARS-CoV-2 vaccination caused monocyte activation and homing, reflected by higher expression of activation markers (CD40, TLR-4, TLR-2 and CD49d) and lower percentage of CCR2, CD11b and CX3CR1 expressing intermediate and non-classical monocytes after vaccination. It has been described, that in addition to DCs, these monocyte subsets also migrate from bloodstream to lungs in patients with COVID-19 (Sánchez-Cerrillo et al., 2020). However, vaccine-induced monocyte homing was found mainly in young people. As same as we observed in CD1c+ mDCs, less monocyte homing was associated to a lower specific T cell response to vaccination. Although monocytes have not a main role in the modulation of T cell response, monocytes are known to have the ability to prime tissue resident T cells via cytokine production (Thompson et al., 2019). Thus, a deficit in monocyte migration to inflammatory sites might also negatively affect T cell response.

One of the key age-related immune defects is the phenomenon called inflammaging, a persistent increase in basal pro-inflammatory phenotype found in the elderly (Franceschi et al., 2017). Monocytes are one of the principal players of inflammaging (De Maeyer and Chambers, 2021; Shaw et al., 2010). Inflammation is a critical factor in COVID-19 progression where monocyte-driven cytokine storm induces a hyper-inflammatory phenotype leading to a more severe symptomatology in COVID-19 patients (Vanderbeke et al., 2021). Here, we discovered a higher TLR-4-mediated pro-inflammatory cytokine production by monocytes from elderly people after SARS-CoV-2 vaccination and importantly, this cytokine production was inversely correlated to specific T cell response. This is in accordance to previously published studies, describing the role of inflammatory monocytes in the suppression of vaccine responses (Mitchell et al., 2012; Moyat et al., 2015). In fact, increased gene expression of inflammatory responses and TNF-α signaling via NFκβ were reported after COVID-19 vaccination (Liu et al., 2021). Importantly, plasma TNF-α levels in old people were associated to a poorer antibody response following SARS-CoV-2 vaccination (Demaret et al., 2021). According with our results from monocytes, old people showed increased numbers of CD16+ mDCs, a DC subset that also participate in inflammatory responses (Piccioli et al., 2007). This higher CD16+ mDC levels were inversely associated to SARS-CoV-2 specific T cell response after vaccination. Altogether, a higher capacity of monocytes to produce pro-inflammatory cytokines, increased CD16+ mDC numbers and even the higher T cell activation status found in elderly people are probably associated to the inflammaging phenomenon, which at the same time is related to lower vaccine immunity.

In conclusion, we describe age-related innate and adaptive immune deficits associated to a lower SARS-CoV-2 vaccination response, including thymic dysfunction and subsequent alteration of T cell homeostasis, an impaired DC homing and function and monocyte-mediated pro-inflammatory profile. These findings contribute to a better understanding of why elderly people are less capable to respond to SARS-CoV-2 vaccination and might be relevant for the improvement of the current vaccination strategies, especially in this vulnerable population, and for the development of more efficient prototypes for the general population.

## LIMITATIONS OF THE STUDY

One of the limitations of the present study might be the sample size. However, we performed an exhaustive and comprehensive analysis of age-associated immune deficits and the differences and associations among the contrasts were very clear. Additionally, it may be interesting to carry out the same determinations long-term after SARS-CoV-2 vaccination. Nevertheless, the described age-associated immune defects are already observed two months after the second dose, therefore, these defects are expected to be maintained or even increase with a longer follow up. Lastly, another limitation of this work could be that we only studied the response to BNT162b2 mRNA vaccine. Nevertheless, it is one of the most administered COVID-19 vaccines and it would not be surprising that other vaccines present similar results, especially other mRNA vaccines.

## Data Availability

All data produced in the present study are available upon reasonable request to the authors

## ACKNOWLEDGMENTS

We thank all the community volunteers, participants from the Institute of Biomedicine of Seville and nursing staff from Virgen del Rocío University Hospital who have taken part in this project. We also thank Alicia Gutierrez and Esperanza Muñoz for reviewing the manuscript. This work was supported by Consejería de Transformación Económica, Industria, Conocimiento y Universidades, Junta de Andalucía (CV20-85418 and P20_00906, DOC-01659 and DOC-00963); Consejería de Salud, Junta de Andalucía (RH-0037-2020), Instituto de Salud Carlos III (CP19/00159, FI17/00186, FI19/00083, PI19/01172, CM20/00243) Fondos FEDER, and National Spanish Research Council (CSIC).

## AUTHOR CONTRIBUTIONS

E.R.M. conceived and designed the research. A.P.G., C.G.C., I.R.J., M.M.S.S., A.R.M., M.R.I.B. and E.R.M participated in sample collection and processing. J.V., A.P.G., F.J.O., C.G.C. and M.R.J.L performed the experiments. J.V. carried out the data analysis. J.V., S.B., L.F.L.C., M.R.I.B. and E.R.M. participated in the interpretation/discussion of the results. J.V., A.P.G. and E.R.M wrote the manuscript. E.R.M. and J.V. coordinated the project. All authors reviewed and discussed the manuscript.

## DECLARATION OF INTERESTS

The authors declare no competing interests.

## MATERIAL AND METHODS

### HUMAN SUBJECTS / STUDY PARTICIPANTS

Fifty-four participants vaccinated with BNT162b2 mRNA vaccine against SARS-CoV-2 were included in this study. They were recruited at the Institute of Biomedicine of Seville and community volunteers from Seville, Dos Hermanas (Seville, Spain) and Rota (Cadiz, Spain). Inclusion criteria included subjects with self-sufficient health status and participants were excluded if they had a diagnose of dementia, active infections or hospital admission during the last six months. Three young subjects were excluded from the study due to a positive result for SARS-CoV-2 PCR or SARS-CoV-2 RBD-specific antibodies prior to vaccination. Participants were stratified by age: <60 categorized as young (n=33) and ≥60 categorized as old (n=21) the median ages of young and old subjects were 29 [IQR, 26-49] and 73 [72-74], respectively. Peripheral blood samples were extracted from February to November 2021.

### STUDY DESIGN

A longitudinal design was performed to study the immune parameters associated to vaccination response to BNT162b2 vaccine against SARS-CoV-2 between old and young people. Three time-points were studied to analyze the innate and adaptive immunity parameters: (1) before the administration of the first dose (PRE); (2) three weeks after the first dose, just before the administration of the second dose (1D); and, (3) two months after the second dose (2D) (Figure S1). Humoral and cellular responses were studied. First, SARS-CoV-2 specific response was measured by the quantification of IgG levels against SARS-CoV-2 S1 receptor binding domain (RBD) by enzyme-linked immunosorbent assay (ELISA). Then, the spike-specific T cell response was analyzed using flow cytometry. T lymphocytes, dendritic cells (DCs) and monocytes were phenotypically analyzed by multiparametric flow cytometry. TLR-dependent function was also measured in myeloid dendritic cells and monocytes using flow cytometry, and plasmacytoid dendritic cells (pDCs) by ELISA. All these immune parameters were correlated with SARS-CoV-2 specific humoral and T cell responses after the vaccination.

### ETHICS STATEMENT

Participants gave written consent and the study was approved by the Ethics Committee of the Virgen del Rocio University Hospital (protocol code “COVIMARATON_2020”; internal code 0156-N-21).

## METHODS DETAILS

### CELL AND PLASMA ISOLATION

Peripheral blood mononuclear cells (PBMCs) and plasma were isolated from study subjects’ blood. PBMCs were isolated using BD Vacutainer® CPT™ Mononuclear Cell Preparation Tubes (with Sodium Heparin, BD Biosciences) in a density gradient centrifugation at the same day of blood collection. CPTs were centrifuged at 3000 rpm for 20 min at room temperature. Afterwards, PBMCs were cryopreserved in freezing medium (90% of fetal bovine serum (FBS) (Gibco) + 10% dimethyl sulfoxide (DMSO) (PanReac AppliChem) in liquid nitrogen until further use. Plasma samples were obtained using BD Vacutainer™ PET EDTA Tubes centrifugation at 3000 rpm for 5 min, aliquoted and cryopreserved at −80°C until further use.

### CELL PREPARATION AND STIMULATION

PBMCs were thawed, washed using RPMI 1640 (Gibco) and rested for 1 h in 0.25 µL/mL DNase I (Roche Diagnostics) containing R-10 complete medium (RPMI 1640 supplemented with 10% FBS, 100 U/ml penicillin G, 100 l/ml streptomycin sulfate (Gibco), and 1.7 mM sodium L-glutamine (Lonza)).

#### SARS-CoV-2 specific T cell response

To analyze the specific T cell response to SARS-CoV-2, 1.5 × 10^6^ PBMCs were *in vitro* stimulated for 6 h at 37°C in R-10 medium with overlapping peptides of protein Spike (PepMix™ SARS-CoV-2; Spike Glycoprotein, from JPT). 1.5 × 10^6^ PBMCs incubated with the proportional amount of DMSO were included in each batch of experiments as a negative control. The stimulation was performed in the presence of 10 µg/mL of brefeldin A (Sigma Chemical Co) and 0.7 µg/mL of monensin (Golgi Stop, BD Biosciences) protein transport inhibitors, anti-CD107a-BV650 (clone H4A3; BD Biosciences) monoclonal antibody and purified CD28 (clone CD28.2) and CD49d (clone 9F10) (BD Biosciences) as previously described (Ferrando-Martinez et al., 2012; Pérez-Gómez et al., 2021). Intracellular cytokines and cytotoxicity markers were analyzed by multiparametric flow cytometry. Specific T cell response was defined as the frequency of cells expressing intracellular cytokines and/or cytotoxicity markers after the stimulation with Spike peptides minus the levels of this response in the unstimulated condition (background subtraction).

#### Monocytes stimulation

1 × 10^6^ PBMCs were *in vitro* stimulated in a 48-well plate for 5 h at 37°C with 0.5 µL/ml of lipopolysaccharide (LPS, Invivogen) in R-10 medium, including 1 × 10^6^ PBMCs without any stimulation as a negative control. 0.7 µg/mL of monensin (Golgi Stop, BD Biosciences) was added to all experimental conditions. Intracellular cytokines were analyzed by flow cytometry.

#### Myeloid Dendritic Cells stimulation

0.5 × 10^6^ PBMCs were *in vitro* stimulated in a 24-well plate for 24 h at 37°C with 2 µL/ml of polyinosinic:polycytidylic acid (Poly-I:C, InvivoGen) in R-10 medium. 0.5 × 10^6^ PBMCs incubated without stimulus were included as a negative control. Surface expression of activation markers were analyzed by flow cytometry.

#### pDCs stimulation culture and quantification of IFN-α production

0.5 x 10^6^ thawed PBMCs were incubated at 37°C and 5% CO_2_ during 18 hours in R-10 medium with or without 1 µM of the TLR9 agonist CpG-A (InvivoGen). After incubation, cells were pelleted and the supernatants conserved at −80°C for the subsequent quantification of interferon (IFN)-α production by ELISA according manufacturer’s instructions (PBL Interferon Source).

### MULTIPARAMETRIC FLOW CYTOMETRY

In general, for *ex vivo* phenotyping and functional assays, PBMCs were washed (1800 rpm, 5 min, room temperature) with Phosphate-buffered saline (PBS (Gibco)). PBMCs were then incubated 35 min at room temperature (RT) with a viability marker (LIVE/DEAD Fixable Aqua or Violet Dead Cell Stain; Life Technologies) and all the extracellular antibodies (see below). PBMCs were washed and fixed and permeabilized with BD Cytofix/CytoPerm (BD Biosciences) at 4°C for 20 min or Fixation/Permeabilization Buffer Set (eBioscience) at 4°C for 45 min following the manufacturer’s protocol. Then, cells were stained at 4°C for 30 min with intracellular antibodies (see below) and washed. Finally, cells were fixed for 20 minutes at 4°C with 4 % paraformaldehyde solution (PFA (Sigma-Adrich)).

To assay T cell specific response, PBMCs were extracellularly stained with LIVE/DEAD Fixable Aqua Dead Cell Stain (Life Technologies), anti-DUMP-channel-BV510 (CD14-clone MφP9, CD19 clone SJ25C1, CD56 clone NMCAM16.2), anti-CD8-APC (clone SK-1), anti-CD3-BV711 (clone SP34-2), anti-CD45RA-FITC (clone L48), anti-CD27-APCH7 (clone M-T271) anti-PD-1-BV786 (CD279, clone EH12-1) (BD Bioscience) and anti-TIGIT-PerCPCy5.5 (clone A15153G) and anti-LAG3-BV605 (clone 11C3C65) (BioLegend). They were permeabilized and fixed with Cytofix/CytoPerm buffer (BD Bioscience). Cells were intracellularly stained with: anti-IL-2-BV421 (clone MQ1-17H12), anti-IFN-γ-PE-Cy7 (clone B27) (BD Bioscience), anti-TNF-α-AF700 (clone Mab11) (BD Pharmingen) and anti-Perforin-PE (clone B-D48) (BioLegend). For T cell phenotyping, cells were extracellularly stained with LIVE/DEAD Fixable Aqua Dead Cell Stain (Life Technologies), anti CD8-PerCP-Cy5.5 (clone SK1), anti-CD45RA-PeCy7 (clone L48), anti-CD3-BV711 (clone SP34-2) (BD Bioscience), anti-HLA-DR-BV570 (clone L243), anti CD161-BV421 (clone HP-3G10) (BioLegend); permeabilized and fixed with Fixation/Permeabilization buffer (eBioscience™); and intracellularly stained with: anti-Ki67 FITC (clone 11F6) (BioLegend). T cells were gated based on the CD3 and CD8 expression. Each subset (Total Memory, Memory; Central Memory, CM; Effector Memory, EM; and terminally differentiated effector memory, TEMRA) was gated based on CD45RA and CD27 expression (Figure S2A).

To assay monocytes functionality, PBMCs were extracellularly stained with LIVE/DEAD Fixable Violet Dead Cell Stain Kit (Life Technologies), anti-DUMP-channel-V450 (CD3 clone SP34-2, CD19 clone HIB19, CD20 clone L27, CD56 clone B159), anti-CD14-BV650 (clone M5E2), anti-CD16-PeCF594 (clone 3G8), anti-HLA-DR-BV570 (clone L243), (BioLegend); permeabilized and fixed with BD Cytofix/CytoPerm (BD Biosciences); and intracellularly stained with anti-IL-6-Pe (clone MQ2-6A3), anti-IL-1α-FITC (clone AS5), anti-TNF-α-AF700 (clone MAb11) (BD Biosciences). To assay monocytes phenotyping *ex vivo*, PBMCs were extracellularly stained with LIVE/DEAD Fixable Violet Dead Cell Stain Kit (Life Technologies), anti-DUMP-channel-V450 (CD3 clone SP34-2, CD19 clone HIB19, CD20 clone L27, CD56 clone B159), anti-CD14-BV650 (clone M5E2), anti-CD16-PeCF594 (clone 3G8), anti-TLR4-BV786 (clone TF901), anti-CD142-Pe (clone HTF-1), anti-CCR5-APC-Cy7 (clone 2D7/CCR5) (BD biosciences), anti-HLA-DR-BV570 (clone L243), anti-TLR2-FITC (clone TL2.1), anti-CD40-APC (clone HB14), anti-CX3CR1-PerCPCy5,5 (clone 2A9-1), anti-CCR2-BV605 (clone K036C2), anti-CD49d-BV711 (clone 9F10) (BioLegend), and anti-CD11b-AF700 (clone VIM12) (Life Technologies, Thermo Fisher Scientific). Monocytes were gated based on the CD14 and HLA-DR markers and non-classical, intermediate and classical subsets were gated based on CD14 and CD16 expression (Figure S5A).

To assay mDC functionality, PBMCs were extracellularly stained with LIVE/DEAD Fixable Aqua Dead Cell Stain (Life Technologies), anti-CD11c-BV650 (clone B-ly6), anti-HLA-DR-BV570 (clone L243), anti-Lin-2-FITC (CD3 clone SK7, CD19 clone SJ25C1, CD20 clone L27, CD14 clone MφP9 and CD56 clone NCAM16.2), anti-CD16-BV605 (clone 3G8) (BD Biosciences), anti-CD1c-APCCy7 (clone L161), anti-CD141-PeCy7 (clone M80), anti-CD86-BV421 (clone 2331 (FUN-1)), anti-CD40-APC (clone HB14) (BioLegend) and anti-CD83-AF700 (clone HB15) (Invitrogen); permeabilized and fixed with BD Cytofix/CytoPerm (BD Biosciences); and intracellularly stained with anti-IDO-Pe (clone eyedio) (eBioscience). For *ex vivo* DC phenotyping, PBMCs were extracellularly stained with LIVE/DEAD Fixable Aqua Dead Cell Stain (Life Technologies), anti-CD11c-BV650 (clone B-ly6), anti-HLA-DR-BV711 (clone G46-6), anti-Lin-2-FITC, anti-CD16-BV605 (clone 3G8), anti-CCR7-BV786 (CD197) (clone 3D12), anti-CD86-BV421 (clone 2331 (FUN-1)), anti-PD-L1 PeCF594 (CD274) (clone MIH1), anti-Integrin-β7-APC (clone FIB504) (BD Biosciences), anti-CD4-PerCPCy5,5 (clone OKT4), anti-CD1c-APCCy7 (clone L161), anti-CD141-PeCy7 (clone M80) (BioLegend) and anti-CD123-AF700 (clone 32703) (R&D Systems) antibodies. Cells were permeabilized and fixed with Fixation/Permeabilization buffer (eBioscience™); and intracellularly stained with anti-IDO-Pe (clone eyeido) (eBioscience). DCs were identified by the expression of HLA-DR and the lack of expression of Lin-2. pDCs and mDCS were gated based on the CD123 and CD11c expression, respectively. mDCs subsets were gated using according to CD16, CD1c and CD141 expression (Figure S4A).

Multiparametric flow cytometry were performed on an LRS Fortessa flow cytometer using FACS Diva software (BD Biosciences), acquiring 0.5-1 × 10^6^ events. Data were analyzed using the FlowJo 10.7.1 software (Treestar, Ashland, OR).

### QUANTIFICATION OF ANTI-RBD IgG LEVELS

Anti-RBD IgG SARS-CoV-2 levels were measured by recombinant (r)RBD specific ELISA as previously described (Grifoni et al., 2020; Rydyznski Moderbacher et al., 2020; Sette and Crotty, 2020; Amanat et al., 2021). Briefly, Nunc Maxisorp flat-bottomed 96-well plates (ThermoFisher Scientific) were coated with 1μg/mL of rRBD protein of the spike (S) antigen of SARS-CoV-2 (Sino Biological, #40592-V08H), overnight at 4°C. The following day, plates were blocked with 3% milk in PBS containing 0.05% Tween-20 for 120 min at RT. Plasma samples were heat inactivated at 56°C for 20 min complement activity. Human plasma samples were diluted at 1:50, 1:100, 1:200, 1:400, or 1:800 in 1% milk containing 0.05% Tween-20 in PBS and incubated for 90 min at room temperature. Plates were washed four times with 0.05% PBS-Tween-20. Human serum standard reference material of anti-SARS-CoV-2 immunoglobulin (first WHO International Standard and International Reference Panel for anti-SARS-CoV-2 immunoglobulin from NIBSC, UK, NIBSC code: 20/150) was used as standard curve to titer anti-SARS-CoV-2 IgG antibody in plasma samples. Human serum standard was added to the plates and serially diluted (twofold dilutions) in 1% milk containing 0.05% Tween-20 in PBS. Pooled plasma samples (NIBSC, UK, NIBSC code: 20/142) obtained from healthy blood donors before 2019 was used as negative control plasma. Secondary antibodies, streptavidin-horseradish peroxidase-conjugated mouse anti-human IgG (Hybridoma Reagent Laboratory, Baltimore, MD,) was used at a 1:5,000 dilution in 1% milk containing 0.05% Tween-20 in PBS. Plates were washed four times with 0.05% PBS-Tween-20. The plates were developed using fast o-Phenylenediamine dihydrochloride Peroxidase Substrate (Sigma-Aldrich), the reaction was stopped using 3 M HCl, and the optical density at 490 nm (OD490) was read on a Multiskan GO Microplate Spectrophotometer (ThermoFisher Scientific) within two hours. Anti-SARS-CoV-2 IgG antibody titers for each donor were calculated as Binding Antibody Units (BAU)/ml according to the manufacturers’ information regarding the WHO Standard and was determined based on sigmoidal dose-response nonlinear regression, 4PL, using GraphPad Prism, version 8.0 (GraphPad Software, Inc.).

### DNA EXTRACTION AND sj/β TREC RATIO MEASUREMENT BY ddPCR

The extraction of the genomic DNA from frozen PBMCs was performed using a blood DNA minikit (Omega; Bio-Tek). The DNA concentration was determined by Qubit assay according to the manufacturer’s protocol (ThermoFisher Scientific).

T cell receptor excision circles (TRECs), DβJβ-TRECs and sj-TREC, were quantified from extracted DNA by droplet digital PCR (ddPCR) (Bio-Rad) based on a previously method (Ferrando-Martínez et al., 2010). 20 µM of primer for DβJβ-TRECs and sj-TREC, 10 µM of probe and ddPCR Supermix for probes no dUTP (Bio-Rad) were used. The reference gene used was RPP30 (two copies per cell). The ddPCR conditions were: 10 min at 95 °C, 40 cycles of 30 sec at 94 °C, 1 min at 59 °C and 10 min at 98°C. The program used for determining the sj/β TREC ratio was Bio-Rad QuantaSoft software v.1.7.4.

## QUANTIFICATION AND STATISTICAL ANALYSIS

### STATISTICAL ANALYSIS

Non-parametric statistical analyses were performed using Statistical Package for the Social Sciences software (SPSS 25.0; SPSS, Inc.), RStudio Version 1.3.959 and GraphPad Prism version 8.0 (GraphPad Software, Inc.). Polyfunctionality pie charts were constructed using Pestle version 1.6.2 and Spice version 6.0 (Roederer et al., 2011). Median and interquartile ranges were used to describe continuous variables and percentages to describe categorical variables. ROUT method was utilized to identify and discard outliers. Differences between old and young groups were analyzed by two-tailed non-parametric Mann-Whitney U test. The non-parametric Wilcoxon test was used to analyze differences between time points. The Spearman test was used to analyze correlations between variables. Permutation test was used to assess differences between pie charts using Spice software. Hmisc and corrplot packages were used in R by Spearman method to calculate correlations between pairs of variables and plot the correlation matrix figures. Lateral intensity bar from red to blue, next to correlation matrixes, represents (Ρ, ρ) rho coefficient value of the Spearman test. All differences with a P value < 0.05 were considered statistically significant (*p < 0.05, **p < 0.01, ***p < 0.001, ****p < 0.0001).

## Notes

### Competing Interest Statement

The authors have declared no competing interest.

### Funding Statement

This study was funded by Consejeria de Transformacion Economica, Industria, Conocimiento y Universidades, Junta de Andalucia (CV20-85418 and P20_00906, DOC-01659 and DOC-00963); Consejeria de Salud, Junta de Andalucia (RH-0037-2020), Instituto de Salud Carlos III (CP19/00159, FI17/00186, FI19/00083, PI19/01172, CM20/00243) Fondos FEDER, and National Spanish Research Council (CSIC).

