## SupplementaryInformation for "Innate and adaptive immune defects associated with lower SARS-CoV-2 BNT162b2 mRNA vaccine response in elderly people"

### Figure S1

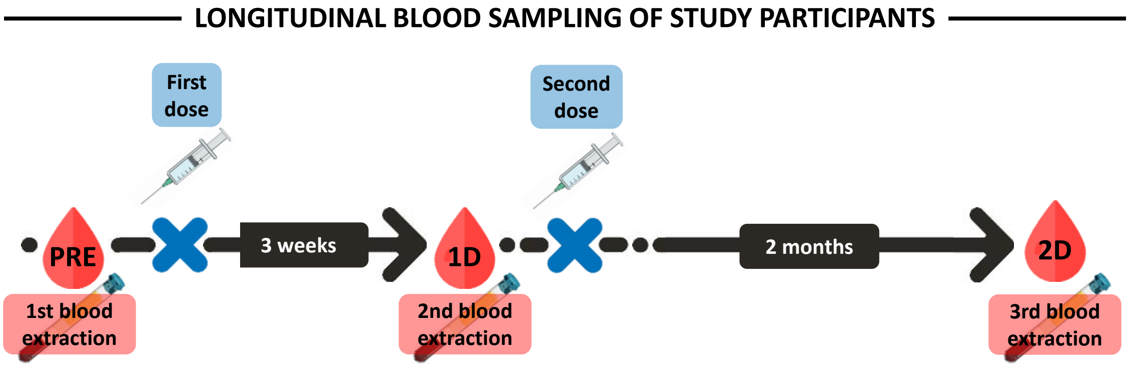


| | All participants (n=54) | Elderly participants (n=21) | Young participants (n=33) |
|---|---|---|---|
| Age (years) | 49.5 [28 – 73] | 73 [72 – 74] | 29 [26 –48.5] |
| Sex (Female sex), n (%) | 35 (64.8) | 13 (61.9) | 22 (66.7) |

### Figure S2

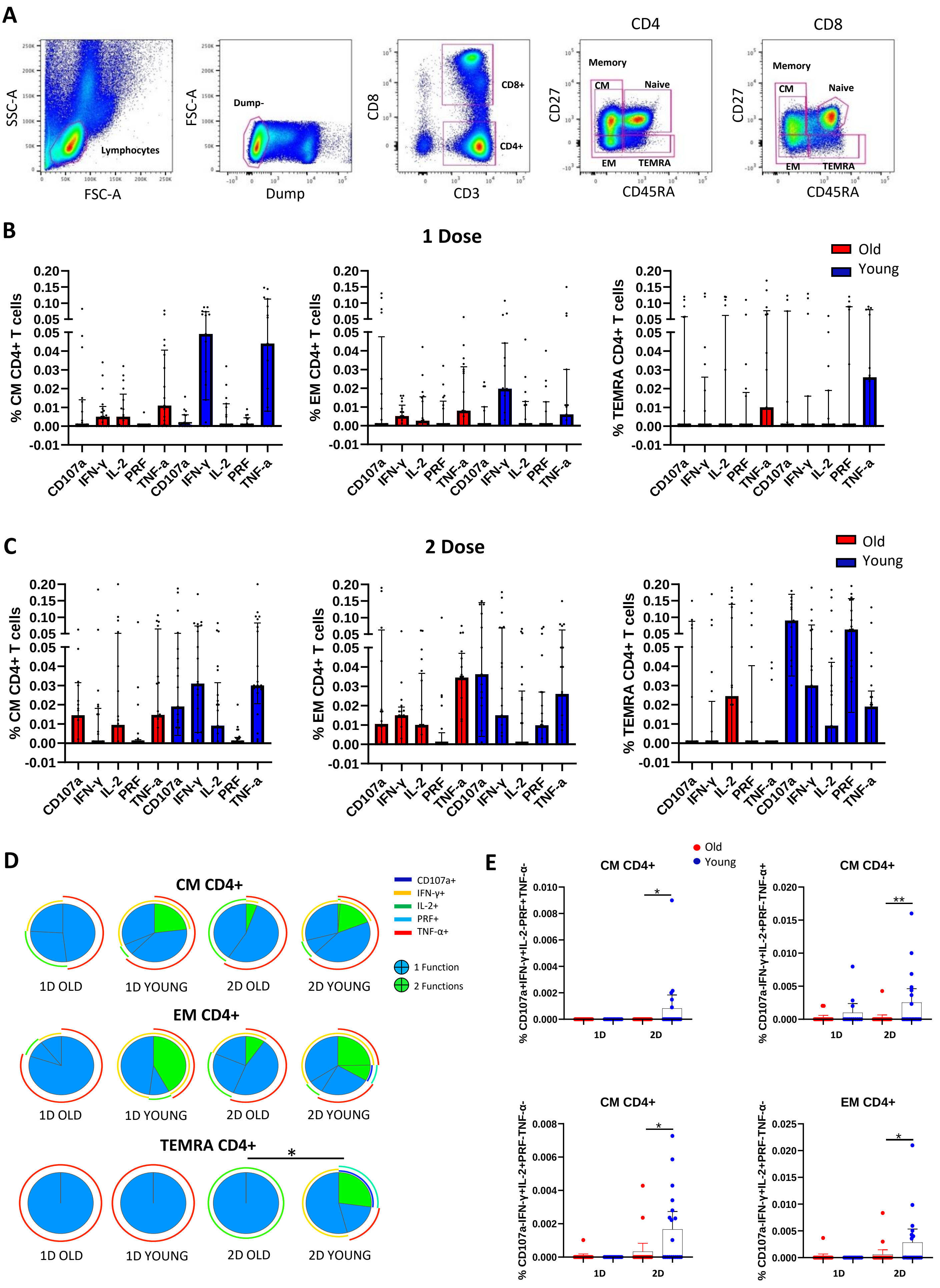

A
SSC-A
FSC-A
Lymphocytes
FSC-A
Dump-
Dump
CD8
CD8+
CD4+
CD3
CD4
CD27
Memory
CM
Naive
EM
TEMRA
CD45RA
CD8
B
1 Dose
Old
Young
% CM CD4+ T cells
% EM CD4+ T cells
% TEMRA CD4+ T cells
CD107a
IFN-γ
IL-2
PRF
TNF-a
C
2 Dose
D
CM CD4+
EM CD4+
TEMRA CD4+
1D OLD
1D YOUNG
2D OLD
2D YOUNG
CD107a+
IFN-γ+
IL-2+
PRF+
TNF-α+
1 Function
2 Functions
E
CM CD4+
EM CD4+
% CD107a+IFN-γ+IL-2-PRF+TNF-α-
% CD107a-IFN-γ+IL-2+PRF-TNF-α+
% CD107a-IFN-γ+IL-2+PRF-TNF-α-
1D
2D

### Figure S3

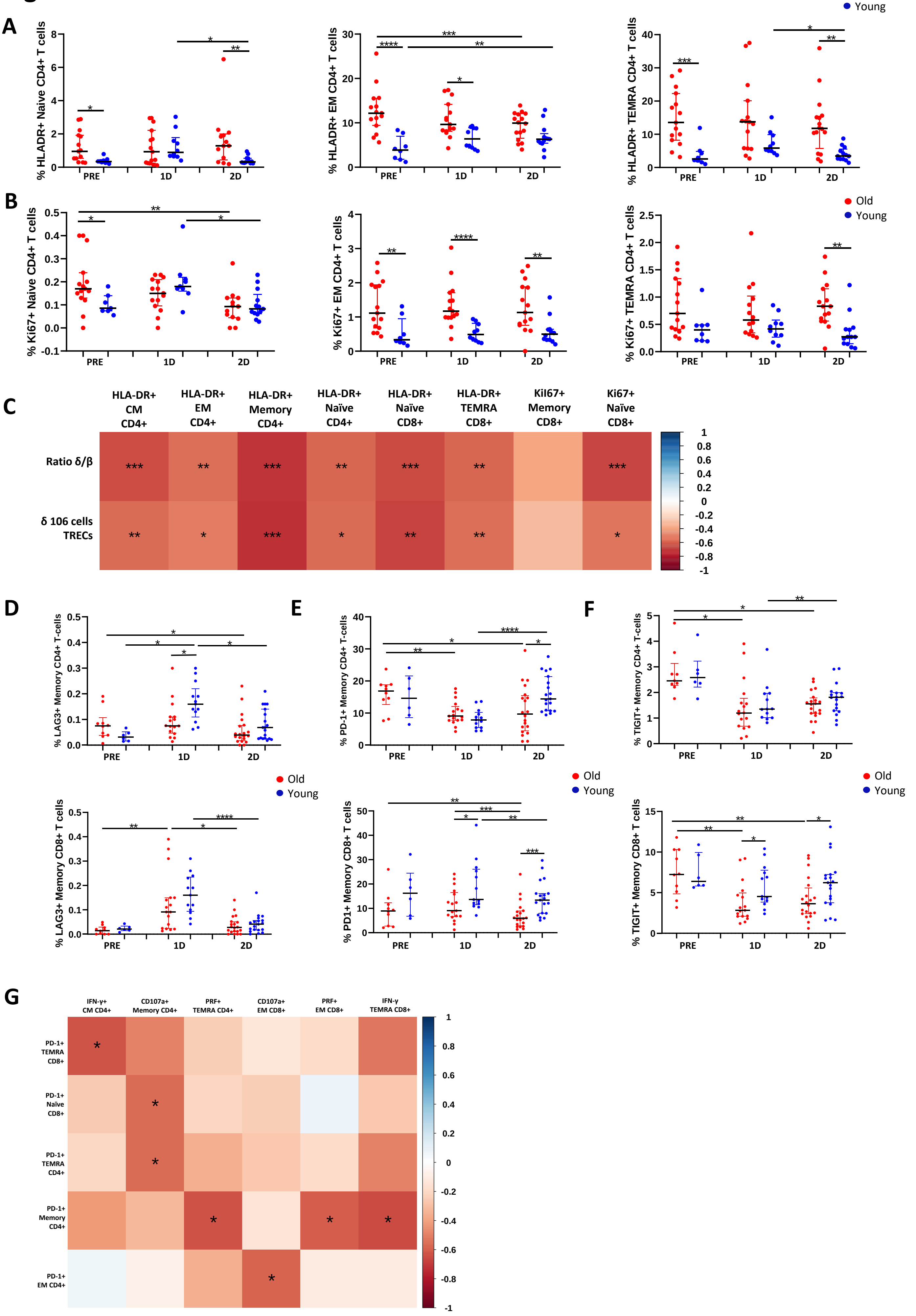
Old
Young
A
% HLADR+ Naive CD4+ T cells
% HLADR+ EM CD4+ T cells
% HLADR+ TEMRA CD4+ T cells
PRE
1D
2D
B
% Ki67+ Naive CD4+ T cells
% Ki67+ EM CD4+ T cells
% Ki67+ TEMRA CD4+ T cells
C
HLA-DR+ CM CD4+
HLA-DR+ EM CD4+
HLA-DR+ Memory CD4+
HLA-DR+ Naïve CD4+
HLA-DR+ Naïve CD8+
HLA-DR+ TEMRA CD8+
Kil67+ Memory CD8+
Ki67+ Naïve CD8+
Ratio δ/β
δ 106 cells TRECs
D
% LAG3+ Memory CD4+ T-cells
% LAG3+ Memory CD8+ T cells
E
% PD-1+ Memory CD4+ T-cells
% PD1+ Memory CD8+ T cells
F
% TIGIT+ Memory CD4+ T-cells
% TIGIT+ Memory CD8+ T cells
G
IFN-γ+ CM CD4+
CD107a+ Memory CD4+
PRF+ TEMRA CD4+
CD107a+ EM CD8+
PRF+ EM CD8+
IFN-γ TEMRA CD8+
PD-1+ TEMRA CD8+
PD-1+ Naïve CD8+
PD-1+ TEMRA CD4+
PD-1+ Memory CD4+
PD-1+ EM CD4+

### Figure S4

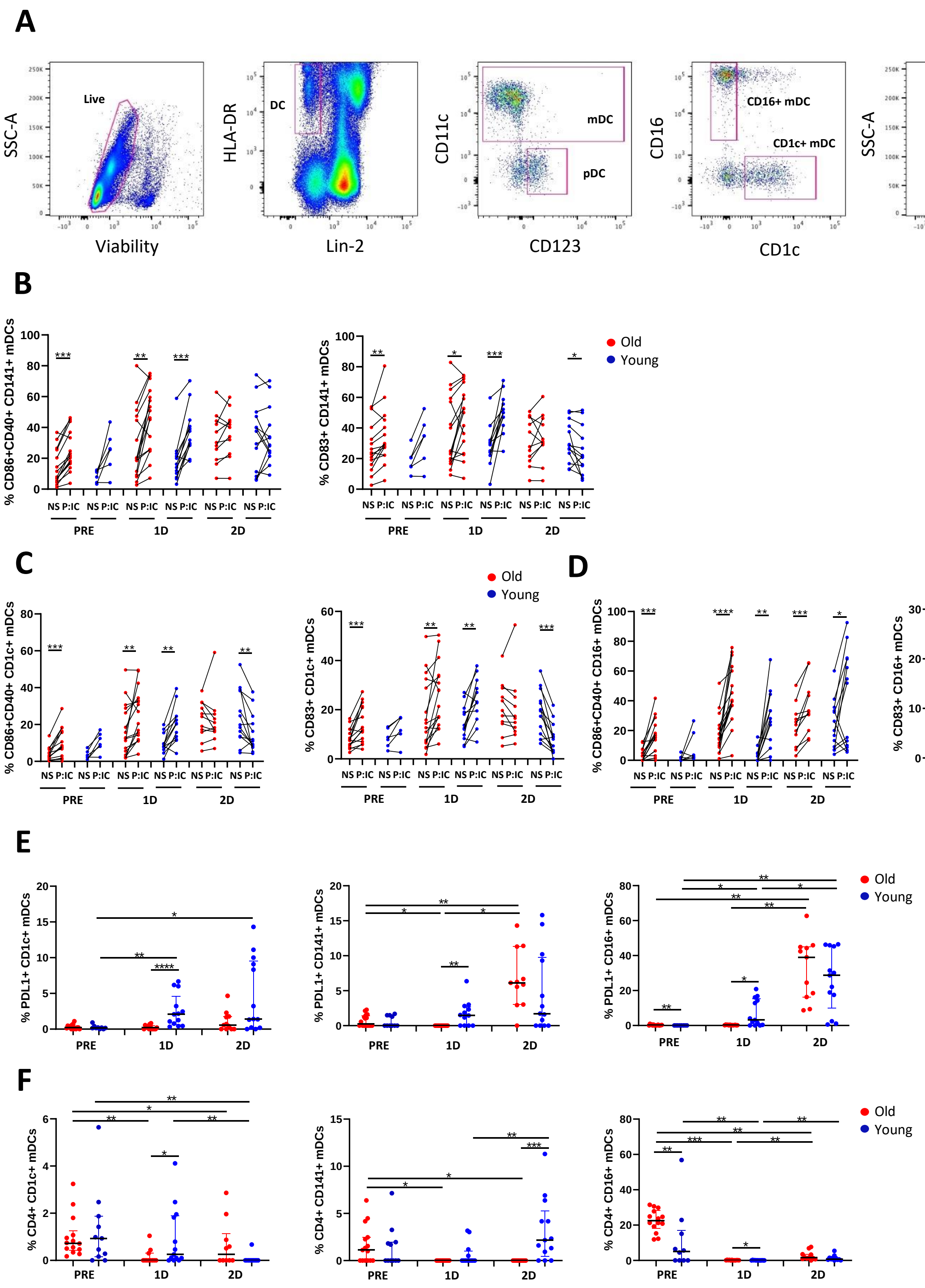

A
SSC-A
Viability
Live
HLA-DR
Lin-2
DC
CD11c
CD123
mDC
pDC
CD16
CD1c
CD16+ mDC
CD1c+ mDC
SSC-A
CD141
CD141+ mDC
B
% CD86+CD40+ CD141+ mDCs
% CD83+ CD141+ mDCs
NS P:IC
PRE
1D
2D
Old
Young
C
% CD86+CD40+ CD1c+ mDCs
% CD83+ CD1c+ mDCs
D
% CD86+CD40+ CD16+ mDCs
% CD83+ CD16+ mDCs
E
% PDL1+ CD1c+ mDCs
% PDL1+ CD141+ mDCs
% PDL1+ CD16+ mDCs
F
% CD4+ CD1c+ mDCs
% CD4+ CD141+ mDCs
% CD4+ CD16+ mDCs

### Figure S5

A

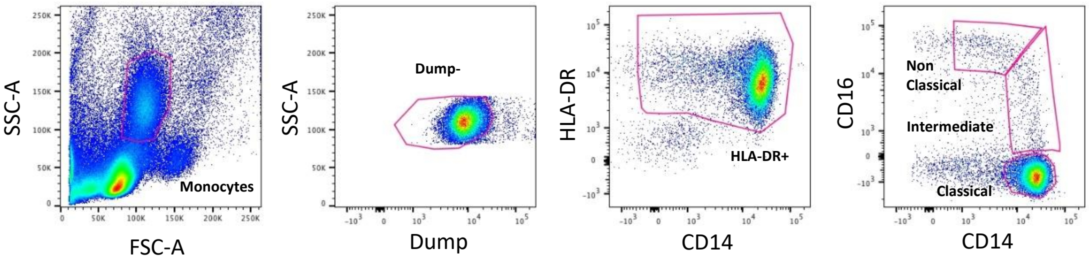


B

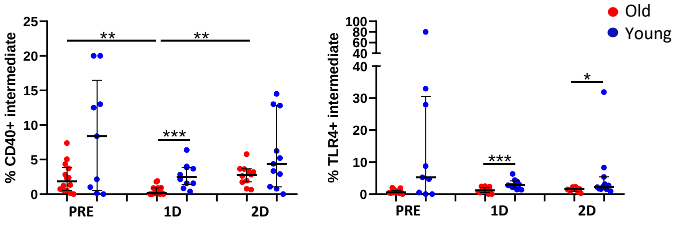


C

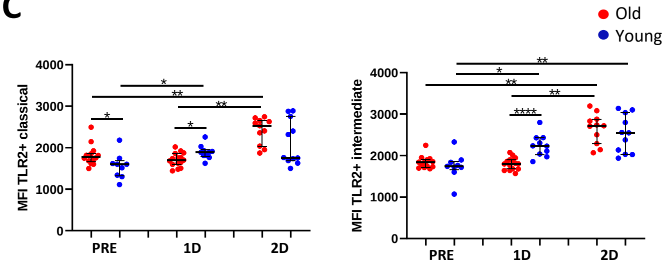


D

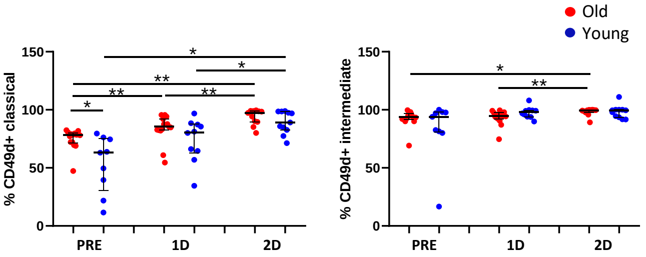


E

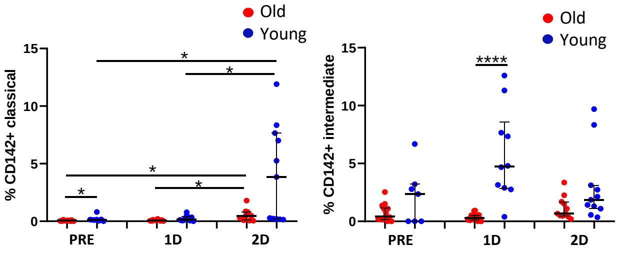

**Figure S1. Flow chart of longitudinal sampling of study participants**

Old and young donors were vaccinated with two doses of BNT162b2 mRNA vaccine, receiving the second dose three weeks after the first one (top). For this study, peripheral blood samples were extracted from all participants just before SARS-CoV-2 vaccination (PRE), three weeks after the first dose and just before the second one (1D), and two months after the second dose (2D) (top). The table describes the age and sex of studied donors (down). Variables are expressed as number (n) and percentages (%), and continuous variables are expressed as median with interquartile ranges [IQR] (down).

**Figure S2. SARS-CoV-2 S-specific T cell response in old and young people**

**(A)** Gating strategy of T cells is shown. Lymphocytes were firstly identified according to their size and complexity (FSC-A and SSC-A) and cells negative for dump channel (viability, CD14, CD19, and CD56) were gated. Then, CD8+ (CD3+ CD8+) and CD4 (CD3+ CD8-) T cells were selected and T cell subsets were identified based on the expression of CD45RA and CD27: naïve (CD45RA+ CD27-) central memory (CM) (CD45RA- CD27+), effector memory (EM) (CD45RA- CD27-) and terminal differentiated effector memory (TEMRA) (CD45RA+ CD27+) cells. Total memory cells (Memory) correspond to the sum of CM, EM and TEMRA T cells.

**(B and C)** Bar graphs showing the percentages of CM, EM and TEMRA CD4+ T cells expressing CD107a, IFN-γ, IL-2, PRF and TNF-α upon S-specific SARS-CoV-2 stimulation, comparing old and young subjects three weeks after the first dose (B) and two months after the second dose (C) of SARS-CoV-2 vaccine.

**(D)** Pie charts representing SARS-CoV-2 S-specific EM, CM and TEMRA CD4+ T cell polyfunctionality. Each sector represents the proportion of S-specific CD4+ T cells producing two (green) or one (blue) functions. Arcs represents the type of function (CD107a, IFN-γ, IL-2, PRF and TNF-α) expressed in each sector.

**(E)** Bar graphs showing the percentage of EM and CM CD4+ T cells expressing different combinations of studied functions (CD107a, IFN-γ, IL-2, PRF and TNF-α) comparing old and young subjects after the first (1D) and the second (2D) dose.

**Figure S3. T cell homeostasis parameters and its association with SARS-CoV-2 specific T cell response in old and young people**

**(A and B)** Bar graphs representing the percentage of naïve (left), EM (middle) and TEMRA (right) CD4+ T cells expressing HLA-DR (A) and Ki67 (B) in old and young participants before SARS-CoV-2 vaccination (PRE), three weeks after the first dose (1D) and two months after the second dose (2D) of vaccination.

**(C)** Correlation matrix representing associations of sj/β-TREC ratio and DβJβ-TREC/$10^6$ cells with the percentages of HLA-DR+ and Ki67+ T cells prior vaccination.

**(D-F)** Bar graphs representing the percentage of memory CD4+ and CD8+ T cells expressing the immune check points LAG-3 (D), PD-1 (E) and TIGIT (F) in old and young participants at the three follow up time points.

**(G)** Correlation matrix representing associations of the percentage of PD-1+ T cells with SARS-CoV-2 S-specific CD4+ and CD8+ T cells expressing IFN-γ and cytotoxicity markers.

**Figure S4. Dendritic cells phenotype and function before and after SARS-CoV-2 vaccination in old and young people**

**(A)** Gating strategy of DCs. First live cells were selected and DCs were identified by gating HLA-DR+ and Lineage-2- cells. Then mDCs (CD11c+) and pDCs (CD123+) were selected and mDC subsets were identified according to the surface expression of CD1c, CD16 and CD141.

**(B-D)** Before and after graphs showing the percentages of CD83+ and CD86+CD40+ CD141+ (B), CD1c+ (C) and CD16+ (D) mDCs without stimulation (NS) or after TLR-3 stimulation with Poly I:C (P:IC) in old and young participants before SARS-CoV-2 vaccination (PRE), three weeks after the first dose (1D) and two months after the second dose (2D) of vaccination.

**(E and F)** Bar graphs showing the percentages of PDL-1+ (E) and CD4+ (F) mDCs in old and young participants at the three follow up time points in the three DC subsets CD1c+, CD141+ and CD16+.

**Figure S5. Monocyte phenotype before and after SARS-CoV-2 vaccination in old and young people**

**(A)** Gating strategy of monocytes. Monocytes were firstly identified according to their size and complexity (FSC-A and SSC-A) and cells negative for dump channel (viability, CD3, CD19, CD20 and CD56) were gated. Then, HLA-DR+ cells were selected and monocyte subsets were identified according to the expression of CD14 and CD16: classical (CD14++ CD16-), intermediate (CD14++ CD16+) and non-classical (CD14+ CD16+).

**(B-E)** Dot plots showing the percentages of CD40+ (left) and TLR-4+ (right) (B), the median fluorescence intensity of TLR-2 and the percentages of CD49d+ and CD142+ monocytes in old and young participants before SARS-CoV-2 vaccination (PRE), three weeks after the first dose (1D) and two months after the second dose (2D) of vaccination.
